# DNA methylation in primary myelofibrosis is partly associated with driver mutations and distinct from other myeloid malignancies

**DOI:** 10.1101/2024.12.02.24318089

**Authors:** Esra Dursun Torlak, Vithurithra Tharmapalan, Kim Kricheldorf, Joelle Schifflers, Madeline Caduc, Martin Zenke, Steffen Koschmieder, Wolfgang Wagner

**Author notes:** **Corresponding author**: Wolfgang Wagner, Helmholtz-Institute for Biomedical Engineering, Institute for Stem Cell Biology, Medical Faculty of RWTH Aachen University, Pauwelsstrasse 20, Aachen, Germany.

## Abstract

Primary myelofibrosis (PMF) is a clonal blood disorder characterized by mutually exclusive driver mutations in *JAK2*, *CALR*, or *MPL* genes. To explore the epigenetic impact of these mutations, we analyzed DNA methylation (DNAm) profiles from PMF patients. Notably, no differences were found in DNAm between *JAK2* and *CALR* mutated cases, whereas *MPL* mutations displayed slightly distinct patterns. Furthermore, induced pluripotent stem cell (iPSC) models with *JAK2* mutations indicated only a moderate association with PMF-related epigenetic changes, suggesting that these alterations may not be directly driven by the mutations themselves. Additionally, PMF-associated epigenetic changes showed minimal correlation with allele burden and were largely influenced by shifts in the cellular composition. PMF DNAm profiles compared with those from other myeloid malignancies - such as acute myeloid leukemia, juvenile myelomonocytic leukemia, and myelodysplastic syndrome – showed numerous overlapping changes, making it difficult to distinguish PMF based on individual CpGs. However, a PMF score created by combining five CpGs was able to discern PMF from other diseases in both training and validation datasets. These findings demonstrate that PMF driver mutations do not directly evoke epigenetic changes. While PMF shares certain epigenetic alterations with other myeloid malignancies, epigenetic signatures can distinguish between PMF and related diseases.

## Introduction

Malignancies are marked not only by genetic mutations but also by epigenetic modifications, though the interplay between the two remains largely unexplored. Myeloproliferative neoplasms (MPNs) offer a unique opportunity to investigate whether specific mutations lead to distinct epigenetic changes, as they feature unique driver mutations in Janus kinase 2 (*JAK2*), particularly at the amino acid position 617 (valine to phenylalanine, V617F), calreticulin (*CALR*), and myeloproliferative leukemia protein (*MPL*) (1). MPN are categorized into various subentities, such as essential thrombocythemia (ET), polycythemia vera (PV) and primary myelofibrosis (PMF), which harbor these mutations at different frequencies (2), with *JAK2* V617F mutations occurring in almost all PV cases but only approximately 50-60% ET and PMF cases, *CALR* mutations occurring in 25-35% of ET and PMF but not PV cases, and *MPL* mutations occurring in 5-8% of ET and PMF but not PV cases (3).

DNA methylation (DNAm) at CG dinucleotides (CpGs) is an epigenetic mechanism that modulates chromatin structure, transcription, and splicing (4). Prior research already demonstrated aberrant DNAm in MPN (5). The different entities of MPN were shown to have similar DNAm changes, which increase during progression and may play an important role in the pathogenesis and leukemic transformation (6). Recently, it has been suggested that DNAm could serve as a biomarker for the fibrotic progression in PMF (7). However, it is so far unclear whether distinct driver mutations are associated with specific epigenetic modifications. Understanding these connections could yield valuable insights into disease mechanisms and identify potential therapeutic targets.

Epigenetic aberrations are also implicated in other myeloid disorders, including acute myeloid leukemia (AML), myelodysplastic syndromes (MDS), and juvenile myelomonocytic leukemia (JMML) (8). However, a comprehensive study that compares these myeloid malignancies with respect to their DNAm profiles is yet elusive. Moreover, many previous studies have not adequately addressed how changes in cellular composition of a given blood sample may affect aberrant DNAm patterns in malignancies (9).

In this study, we have therefore systematically compared the DNAm profiles associated with different driver mutations in PMF and subsequently assessed how these profiles differ from those of other myeloid malignancies.

## Material and Methods

### Blood samples

Peripheral blood mononuclear cells (PBMCs) of 28 patients diagnosed with PMF, 2 patients diagnosed with ET and 10 healthy donors were used for the study. A detailed description of the sample cohort is shown in Supplemental Table S1. All samples were obtained after informed and written consent in accordance with the Declaration of Helsinki and the research was specifically approved by the local ethics committee of RWTH Aachen University (EK 041/15, EK 206/09 and EK 127/12). PBMCs were isolated from peripheral blood via Ficoll density gradient centrifugation and DNA was isolated with the QIAamp DNA Mini Kit (Qiagen, Hilden, Germany).

### Analysis of mutational burden

We employed a clinically validated amplicon-based next-generation sequencing (NGS) panel (Truseq Custom Amplicon Kit, Illumina, San Diego, USA) to analyze the coding regions of 32 genes commonly associated with hematologic malignancies (10). Variants were manually reviewed, applying a bidirectional frequency cutoff of >1% for driver mutations and > 5% for additional mutations.

### Analysis of DNA methylation profiles

Genomic DNA of the above mentioned samples was subjected to bisulfite conversion and analyzed using the Illumina human EPIC methylation microarray version 2 (EPIC v2). We also examined DNAm profiles of induced pluripotent stem cell (iPSC) lines of three PV donors that are either WT, homozygous or heterozygous for the *JAK2* V617F mutation (supplemental methods). iPSC clones and their iPSC-derived hematopoietic progenitors (iHPCs) were analyzed with EPIC version 1 (EPIC v1). Additionally, we included DNAm profiles of an earlier study on 4 PV, 4 MF, and 4 healthy control samples hybridized on the 450k BeadChip for our validation cohort. All Illumina BeadChip microarrays were analyzed at Life and Brain (Bonn, Germany). Further details on preprocessing and DNAm analysis are provided in the supplemental methods.

To mitigate the impact of variations in cellular composition, we retrieved 289 DNAm profiles of sorted human hematopoietic cell types from the GEO database, including B cells (n=60), CD4 T cells (n=63), CD8 T cells (n=56), granulocytes (n=34), monocytes (n=61), NK cells (n=6), and dendritic cells (n=9; Supplemental Table S2). Pairwise comparisons of mean DNAm values across all cell types were conducted, focusing on CpGs that did not exceed a beta value threshold of 0.1 in any of these comparisons (393,675 CpGs). For further analysis, we concentrated on CpGs that were also detected in all datasets of myeloid malignancies (216,532 CpGs).

For comparison with other myeloid malignancies, we used DNAm datasets of MDS (GSE221745: n=5; GSE152710: n = 73), JMML (GSE237299: n=41), and AML (GSE212937: n=5; GSE62298: n=68), as well as, healthy control datasets of either peripheral blood (GSE141682: n=42; GSE221745: n=5) or bone marrow (GSE221745: n=7; GSE124413: n=40; GSE152710: n=10 samples). For validation, we used available dataset of PMF (GSE152519: n=35; GSE118241: n=22), secondary MF (GSE118241: n=17), ET (GSE156546: n=32), AML (GSE159907: n=316), pediatric AML (GSE133986: n=64) and healthy controls (GSE118241: n=6; GSE156546: n=2; Supplemental Table S3).

### Additional methods

Additional methods for preprocessing and further analysis of DNAm profiles, correlation with gene expression data, generation and characterization of hematopoietic differentiation of iPSC lines, targeted bisulfite amplicon sequencing, and colony-forming unit (CFU) assays are detailed in the supplemental methods.

## Results

### Aberrant DNA methylation in primary myelofibrosis

We examined DNAm profiles of peripheral blood mononuclear cells of PMF patients with *JAK2* V617F mutation (n = 10), *CALR* mutation (n = 10), *MPL* mutation (n = 10, including two ET samples), alongside healthy controls (n = 10; Supplemental Table S1). Multidimensional scaling plots revealed distinct separation between DNAm profiles of PMF and healthy samples, without clear difference based on specific driver mutations (Figure 1a). While samples with additional mutations tended to cluster further away from controls, they did not exhibit a clear separation of specific mutations.

**Figure 1:**
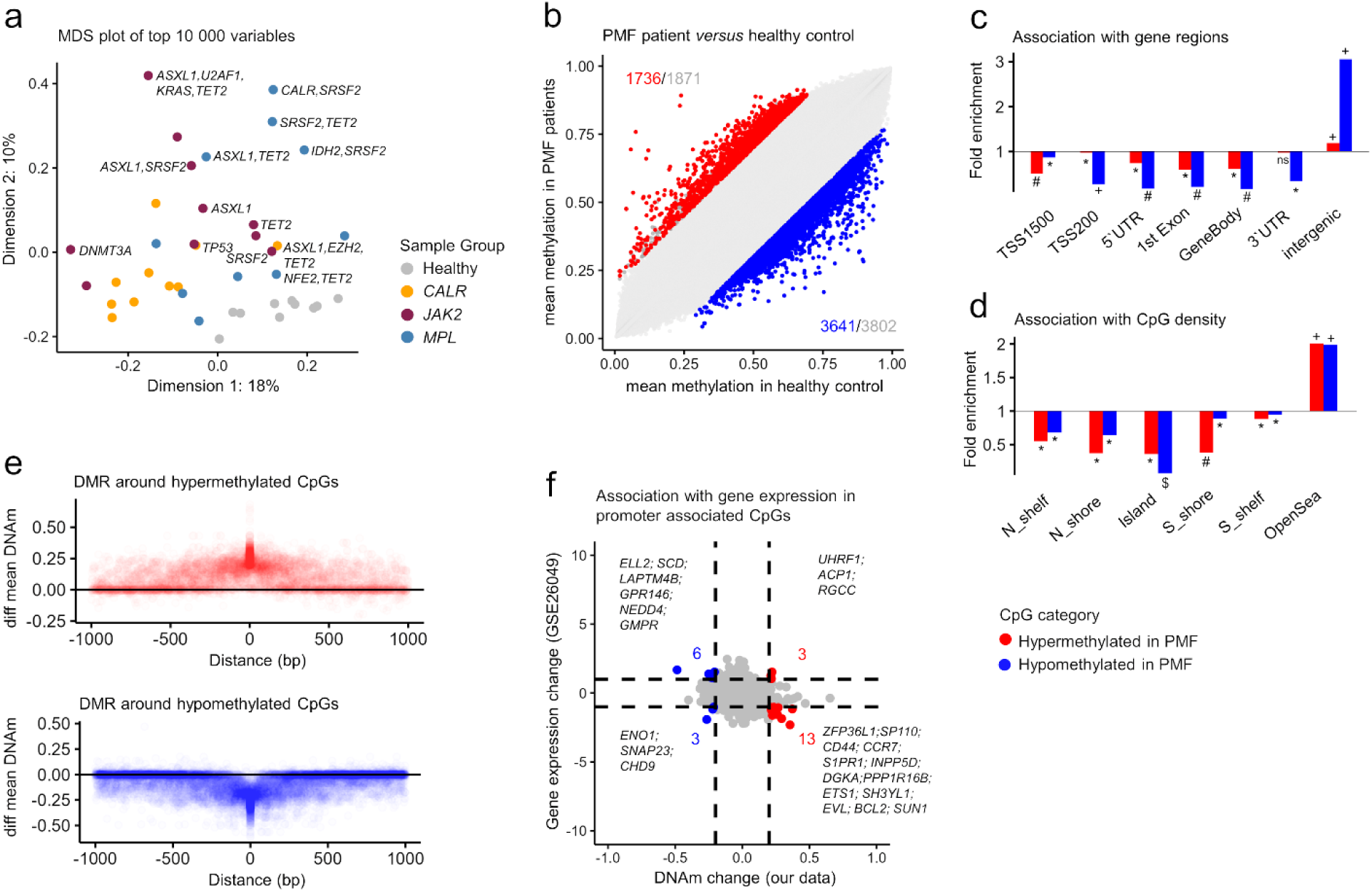
Aberrant DNA methylation in primary myelofibrosis. **a)** Multidimensional scaling plot of DNA methylation profiles in PMF patients with different driver mutations (*JAK2*, *CALR*, *MPL*) and healthy controls (812,274 CpGs). **b)** Scatter plot of mean methylation beta values of PMF patients and healthy controls. Significant hypo- and hypermethylated CpGs are indicated in blue and red (mean DNAm difference >20%; adjusted p-values < 0.05). Grey numbers indicate all CpGs exceeding the mean DNAm difference >20%, irrespective of statistical significance. **c,d)** Enrichment analysis of significant hyper- and hypomethylated CpGs in PMF patients in c) genomic regions and d) CpG islands (Hypergeometric distribution: * = p < 0.05, # = p < 10^-10^, + = p < 10^-20^, and $ = p < 10^-100^. **e)** Differential mean DNAm of CpGs adjacent to all significant hypermethylated and hypomethylated CpGs (1 kb window). **f)** Comparison of DNA methylation and gene expression changes (GSE26049) between PMF patients and healthy controls, with genes showing significant differences in both categories highlighted.

Comparing PMF (n=28) and control samples, we identified 1,736 hypermethylated and 3,641 hypomethylated CpGs in PMF (adj p < 0.05; difference of mean DNAm > 20%; Figure 1b). The most significantly hypermethylated CpGs were frequently linked to genes involved in the pathogenesis of hematological diseases, including *RUNX1* (11), *BRD4* (12), *SRSF2* (13), *SETBP1* (14) and *TNFSF10* (15). Overall, differentially methylated CpGs were enriched in the Gene Ontology (GO) categories for immune response (Supplemental Figure S1) and were predominantly located in intergenic regions (Figure 1c), rather than with CpG islands commonly associated with promoter regions (Figure 1d).

To assess whether aberrant DNAm affects single CpG sites or broader differentially methylated regions (DMRs), we analyzed DNAm in the surrounding of the 1,736 hyper- and 3,641 hypomethylated CpGs. A significant gain or loss of DNAm was observed within a 500 bp window around these CpGs (Figure 1e). We also compared DNAm changes to gene expression profiles from a publicly available dataset (GSE26049). While no clear overall association was found, some candidate genes exhibited concordant changes in DNAm and gene expression, such as hypermethylated and downregulated *ZFP36L1*, linked to myelofibrosis progression (16) and *INPP5D* known to be downregulated by *JAK2* V617F (17). Conversely, hypomethylated and upregulated genes included *LAPTM4B* (18), and *NEDD4* (19), both associated with oncogenic potential (Figure 1f).

### Similar epigenetic effects from *JAK2* and *CALR* mutations

Next, we categorized PMF samples based on their specific driver mutation. Comparing healthy samples (n=10) with MPN samples harboring specific driver mutations (n=10 each), we identified 2,770 hyper- and 7,611 hypomethylated CpGs for *JAK2*, 2,361 and 9,426 for *CALR,* and 238 and 282, respectively, for *MPL* mutations (all adj p < 0.05; difference of mean DNAm > 20%; Figure 2a-c). Notably, a direct comparison between *JAK2* and *CALR* samples revealed only three significant CpGs, indicating minimal differences in their impact on DNAm profiles (Figure 2d). Comparison of *JAK2 versus MPL* (Figure 2e) and *CALR versus MPL* (Figure 2f) revealed a greater number of significant CpGs (Supplemental Table S4), suggesting that *MPL* mutations have distinct epigenetic consequences compared to *JAK2* or *CALR* mutations (Figure 2g).

**Figure 2:**
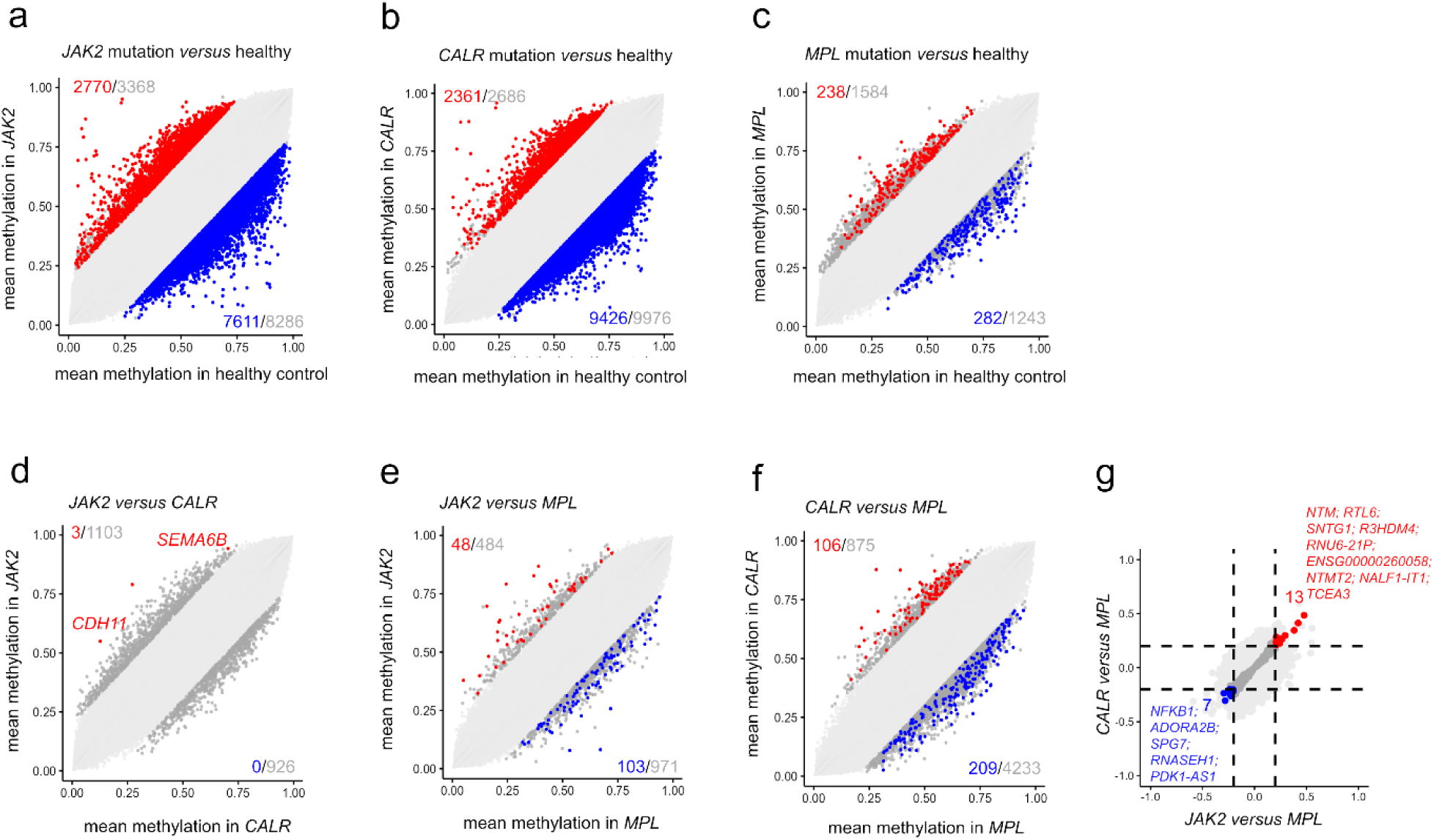
Changes in DNA methylation according to driver mutations. **a-c)** Scatter plots illustrating the mean DNAm beta values in PMF patients compared to healthy controls, stratified by driver mutation: a) *JAK2* mutation, b) *CALR* mutation, and c) *MPL* mutation. **d-f)** Additional scatter plots comparing the mean DNAm beta values of PMF patients based on their driver mutation: d) *CALR versus JAK2;* e) *MPL versus JAK2*; and f) *MPL versus CALR*. Significant hypo- and hypermethylated CpGs are indicated in blue and red, respectively (mean DNAm difference >20%; adjusted p-values < 0.05). **f)** Comparison of significantly differentially methylated CpGs between *JAK2* and *MPL versus CALR* and *MPL*, with gene names for overlapping CpGs highlighted.

### Patient specific DNAm patterns in a *JAK2* iPSC model

To further explore the link between *JAK2* V617F mutations and epigenetic aberrations, we analyzed DNAm profiles in iPSC lines derived from three PV patients previously generated and independent of the above PMF samples, for each of them wild type (WT) *JAK2*, heterozygous (het), and homozygous (hom) *JAK2* V617F mutations (20, 21). The iPSC model offers the advantage of working with clonal and homogenous cell populations, where all cells within a given cell population harbor either heterozygous or homozygous *JAK2* V617F mutations or no *JAK2* mutation. In the pluripotent state, no significant DNAm differences were observed between WT and either het or hom iPSC clones (adj p < 0.05; Figure 3a-b). However, focusing on CpGs exhibiting significant hyper- or hypomethylation in the blood samples of *JAK2* V617F positive patients *versus* healthy controls (2,198 *versus* 5,322 CpGs; lower CpG numbers than in Figure 2a due to the different EPIC array version), we noted a modest yet significant hypomethylation in *JAK2* V617F iPSCs compared to WT iPSCs (Figure 3c-d; Supplemental Figure S2a-b).

**Figure 3:**
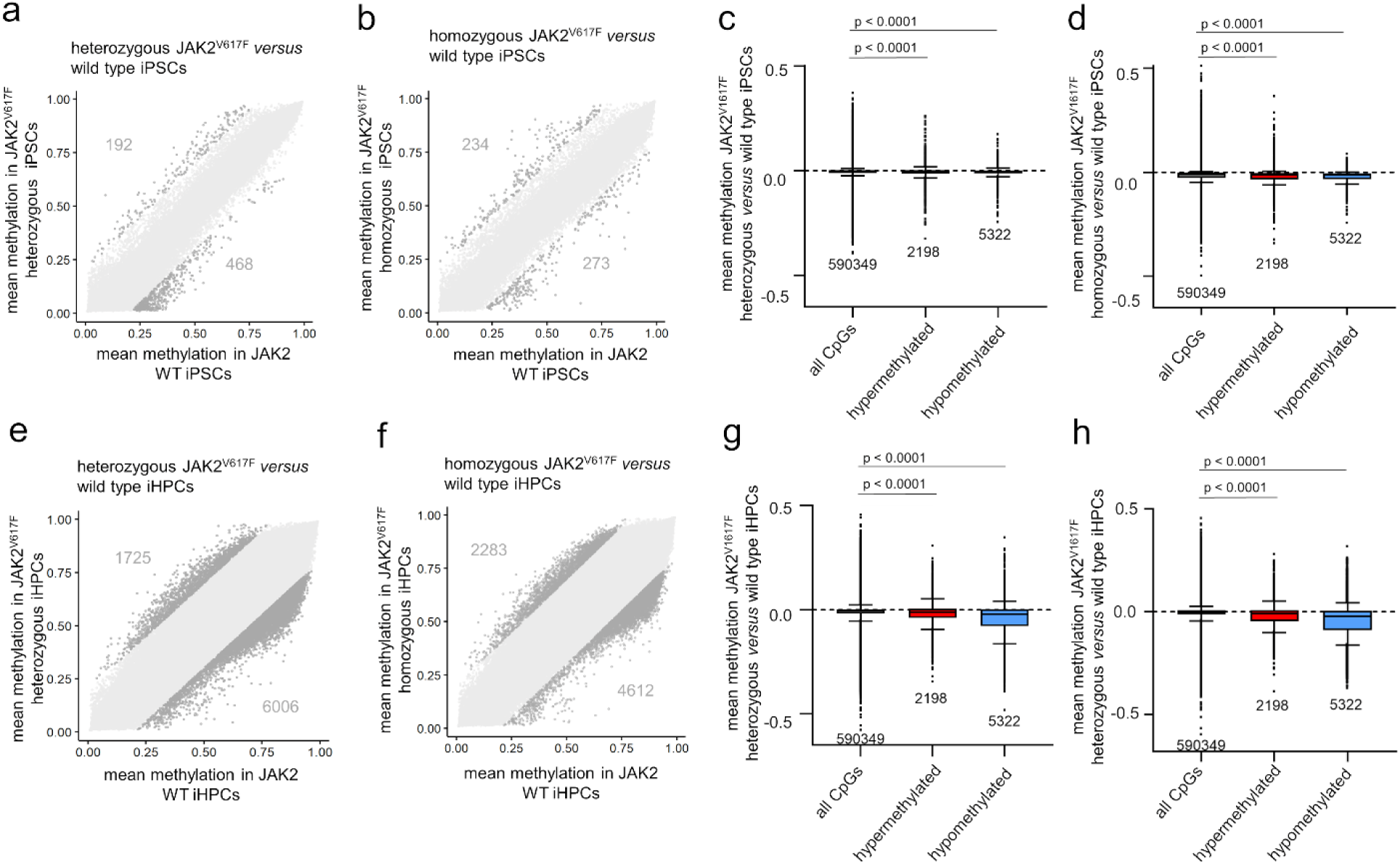
iPSCs with *JAK2* V617F fail to recapitulate disease-associated changes. **a,b)** Scatter plots showing mean DNAm beta values of a) wild type (WT) *versus JAK2* V617F heterozygous (het) iPSCs, and b) WT *versus JAK2* V617F homozygous (hom) iPSCs. The numbers of CpGs with >20% DNAm difference are indicated, but none reached statistical significance. **c,d)** To determine if DNAm changes in PMF patients with *JAK2* V617F are reflected in iPSCs with or without *JAK2* V617F, we focused on CpGs that were significantly differentially methylated in *JAK2* V617F PMF *versus* healthy control (from Figure 2a). Average DNAm changes were then analyzed in these CpGs in iPSCs with either c) WT *versus* heterozygous *JAK2* V617F, or d) WT *versus* homozygous *JAK2* V617F. **e,f)** Following differentiation of iPSC lines into hematopoietic progenitor cells (iHPCs), scatter plots depict mean DNAm beta values for e) WT *versus JAK2* V617F heterozygous iHPCs, and f) WT *versus JAK2* V617F homozygous iHPCs (none of the CpGs reached statistical significance). Grey numbers indicate all CpGs exceeding the mean DNAm difference >20%, irrespective of statistical significance. **g,h)** The CpGs with significant differences in *JAK2* V617F PMF *versus* healthy controls were reanalyzed in iHPCs: g) heterozygous and h) homozygous *JAK2* V617F iHPCs exhibited an overall decrease in DNAm at CpGs that gained or lost methylation in *JAK2* V617F PMF. Statistical significance was evaluated using one-way ANOVA.

To determine whether the impact of *JAK2* V617F mutations was masked in the pluripotent state, we differentiated these clones into hematopoietic progenitor cells (iHPCs; Supplemental Figure S3a). After 16 days, all clones generated non-adherent cells exhibiting typical hematological morphology, with flow cytometry confirming upregulation of various hematopoietic markers (Supplemental Figure S3b-c). Heterozygous and homozygous *JAK2* V617F iHPCs showed a bias towards CD235a/glycophorin A positive erythroid cells in line with our previous study (20). DNAm profiles validated that iHPCs had exited the pluripotent state and were aligned towards mesodermal lineage (Supplemental Figure S3d-e) (22). When comparing iHPCs to iPSCs, we identified 3,201 hypermethylated and 25,507 hypomethylated CpGs (adj p < 0.05; difference of mean DNAm > 20%; Supplemental Figure S3f). These CpGs were enriched in the Gene Ontology categories including blood vessel development, regulation of signaling and migration, indicating that the differentiation captured epigenetic changes of hematopoietic development (Supplemental Figure S3g). However, even in iHPCs there were no significant differences between WT and *JAK2* V617F mutated clones. This lack of distinction might stem from inherent variability during the differentiation process (Figure 3e-f).

Focusing on the CpGs with notable DNAm changes in *JAK2* V617F patients, we found that hypomethylated CpGs in *JAK2* V617F PMF also exhibited hypomethylation in iHPCs carrying the *JAK2* V617F mutation. However, also the hypermethylated CpGs in patients showed moderate hypomethylation in iHPCs (Figure 3g-h). A direct comparison of DNAm changes associated with *JAK2* V617F in iHPCs and PMF patients revealed a moderate but significant association between hypomethylated regions (Supplemental Figure S2c-d). This could be linked to a partial recapitulation of MPN phenotype observed in the *JAK2* V617F iPSC clones. Overall, our findings suggest that the iPSC model does not accurately reflect the DNA methylation changes seen in PMF, indicating that the epigenetic alterations may not be directly driven by the *JAK2* V617F mutation alone.

### Epigenetic age is accelerated in primary myelofibrosis

Subsequently, we investigated if the acceleration of epigenetic age predictions in PMF varied among samples with different driver mutations. Building on our earlier research using bisulfite amplicon sequencing (BA-seq) of three age-associated regions of *PDE4C, FHL2*, and *CCDC102B*, which indicated an overall acceleration of epigenetic age in MPN (23), we employed two epigenetic signatures (24, 25) to further validate that PMF exhibits significantly accelerated epigenetic age, consistent across all three driver mutations (Figure 4a-d).

**Figure 4:**
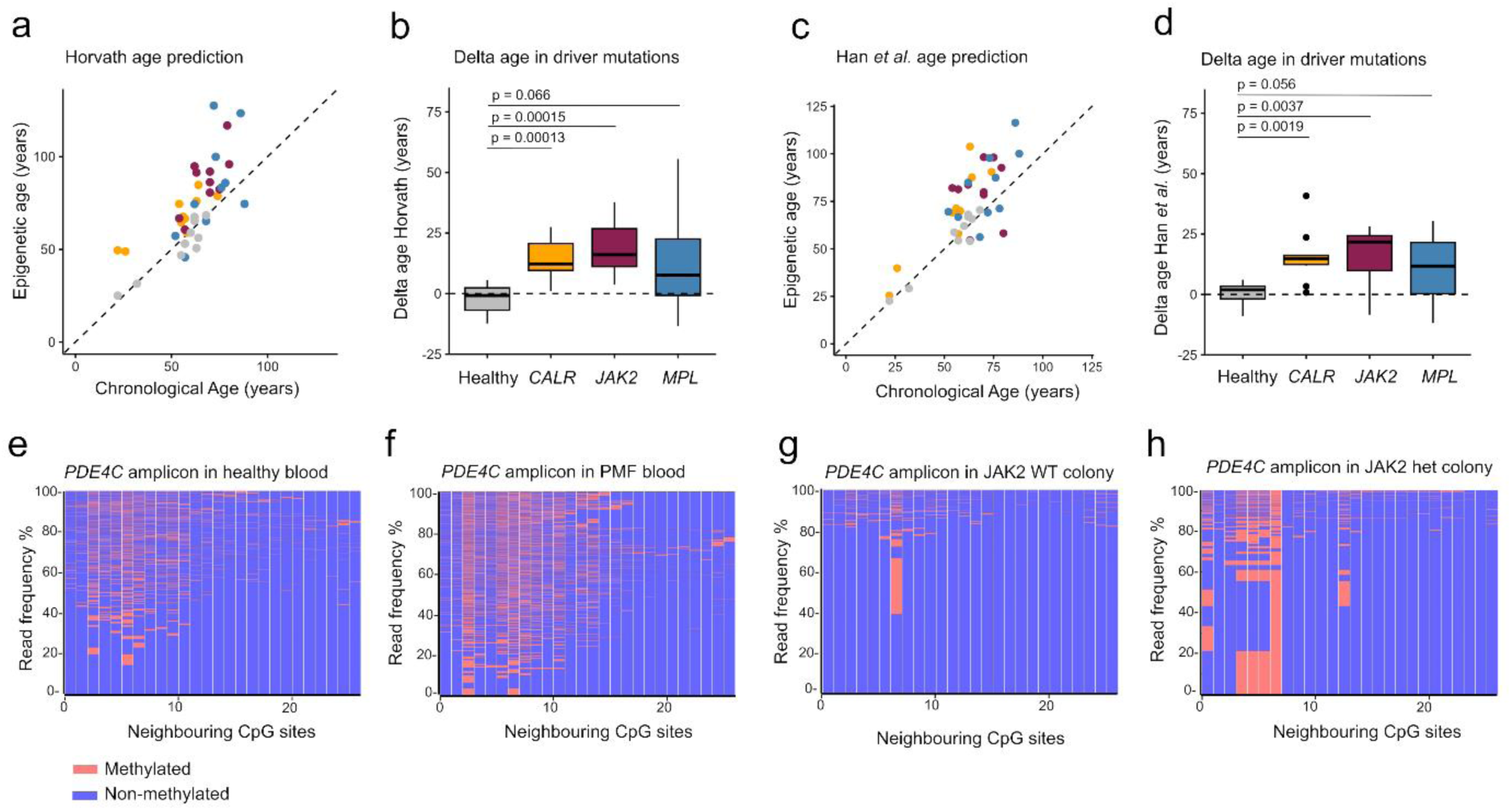
Age-associated DNAm changes in primary myelofibrosis. **a-d)** The correlation between epigenetic age predictions with chronological age and the difference between predicted and chronological age (delta-age) was determined with epigenetic clocks developed by a,b) Horvath (25) and c,d) Han et al. (24). Statistical significance was determined using an unpaired t-test. **e-h)** The DNAm at an age-associated region in *PDE4C* was analyzed by bisulfite amplicon sequencing. The heatmaps exemplifying frequencies of DNAm patterns in 26 neighboring CpGs within the amplicons of e) a healthy donor, and f) a PMF patient blood sample of the same age. The same analysis was performed in colony forming units (CFUs) on day 14 that were either g) a wild-type (WT), or h) harbored the *JAK2* V617F mutation. Unlike the blood samples from PMF patients or controls, the CFUs exhibited a distinct DNAm pattern that appears to reflect the clonal characteristics of the colony-initiating cells.

To delve deeper into the heterogeneity of epigenetic aging, we revisited the BA-seq data of amplicons within the three age-associated regions. Notably, the DNAm at neighboring CpGs within individual reads of amplicons appeared to be independently regulated (Figure 4e-f; Supplemental Figure S4a), corroborating previous findings in healthy samples (24). However, in a clonal disease context, we expected to see a dominant pattern reflective of the tumor-initiating cells. Consequently, we examined BA-seq DNAm patterns at the same age-associated CpGs in single-cell-derived colony-forming units (CFUs; Figure 4g-h; Supplemental Figure S4b). In fact, we observed prominent patterns in CFUs, regardless of whether they are derived from cells with or without *JAK2* V617F mutation, indicating that age-associated DNAm patterns remain largely preserved at least during the CFU formation, possibly influenced later by stochastic factors.

### Disease-associated DNAm and cellular composition

We hypothesized that the allele burden of driver mutations serves as a proxy for the fraction of malignant cells, potentially correlating with PMF-associated aberrant DNAm. In blood samples from PMF patients, allele frequencies ranged from 26% to 93%, although these values were likely different in the PBMCs due to depletion of granulocytes. Multidimensional scaling analysis revealed minimal clustering of DNAm profiles according to allele burden (Figure 5a), suggesting that aberrant DNAm may not be homogeneous across malignant clones. Furthermore, only few individual CpGs exhibited moderate correlation with the allele burden of *JAK2* V617F, *CALR*, or *MPL* mutations (Supplemental Table S5). For example, cg14658896_BC21 (R = 0.62) and cg16965444_BC21 (R = 0.56) displayed an association with allele burden, irrespective of the specific driver mutation (Figure 5b-c).

**Figure 5:**
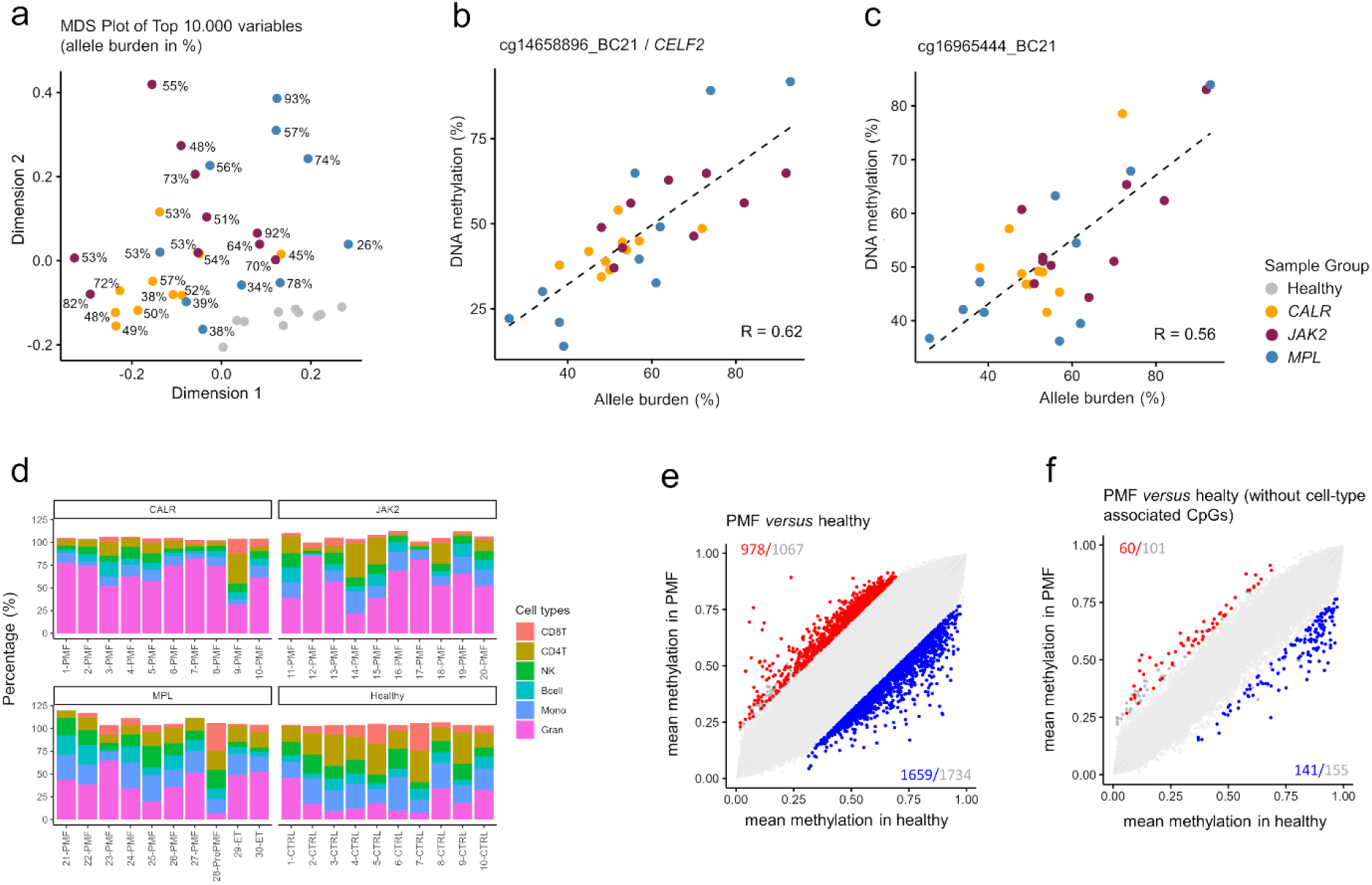
DNA methylation changes are largely attributed to the cellular composition. a) The multidimensional scaling plot demonstrates that PMF sample did not cluster by mutation allele frequency. **b,c)** Correlation between DNAm and allele burden (across all driver mutations, given that they hardly affected DNAm) for the top two candidate CpGs: b) cg14658896_BC21 and c) cg16965444_BC21. **d)** An epigenetic deconvolution algorithm (49) was applied to estimated fractions of granulocytes, monocytes, B cells, NK cells, CD4 and CD8 T cells. **e,f)** To determine how many significant DNAm changes in PMF *versus* controls is attributed to CpGs that have high variation between leukocyte subsets we compared scatter plots e) before and f) after exclusion of CpGs with more than 10% DNAm between any of the leukocyte subsets (only CpGs measured across all datasets are shown). Significant hypo- and hypermethylated CpGs are indicated in blue and red (mean DNAm difference >20%; adjusted p-values < 0.05). Grey numbers indicate all CpGs exceeding the mean DNAm difference >20%, irrespective of statistical significance.

So far, it is largely unclear how much of the aberrant DNAm in myeloid malignancies can be attributed to the changes in the cellular composition of blood. While precise flow cytometric analysis of the leukocyte composition was not available for our samples, we focused on cell type-specific CpGs that exhibited moderate differences between PMF and healthy samples (Supplemental Figure 5) (26–28). Using a deconvolution algorithm for cell type-specific DNAm signatures, we confirmed that the fraction of granulocytes was elevated in PMF compared to healthy controls, with the estimated proportions of different cell types summing close to 100% across samples (Figure 5d).

Given the significant differences in cellular composition between PMF patients and healthy donors, we subsequently concentrated on CpGs exhibiting stable DNAm levels across all healthy leukocyte subsets. This approach was taken to minimize the impact of disparate cellular compositions. Analyzing 289 DNAm profiles from sorted cell types (Supplemental Table S2), we excluded CpGs with over 10% DNAm variation in pairwise comparisons. This process yielded 393,675 CpGs with consistent DNAm across various healthy donor cell types. Comparing PMF and control samples, we found 978 CpGs significantly hypermethylated and 1,659 hypomethylated in PMF (adj p < 0.05; mean DNAm difference > 20%; Figure 5e). However, after excluding CpGs with high-variation between leukocyte subsets, only 60 were significantly hypermethylated and 141 hypomethylated (Figure 5f), indicating that while many PMF-associated epigenetic aberrations are linked to cellular composition, some remain distinctive.

### Comparative epigenetic analysis with other myeloid malignancies

To examine how DNAm profiles in PMF patients compare with other myeloid malignancies, we analyzed datasets from bone marrow samples of myelodysplastic syndrome (MDS) patients (29, 30), peripheral blood of juvenile myelomonocytic leukemia (JMML), and peripheral blood from acute myeloid leukemia (AML) patients (31, 32). To account for potential biases related to tissue type, we compared these against corresponding healthy controls (Supplemental Table S3). Using multidimensional scaling, healthy samples clustered closely together, while JMML samples formed a distinct cluster, and PMF, MDS, and AML profiles did not separate clearly. This observation was very similar either with (Figure 6a-b) or without exclusion of CpGs with high variation between leukocyte subsets (Supplemental Figure 6a-b).

**Figure 6:**
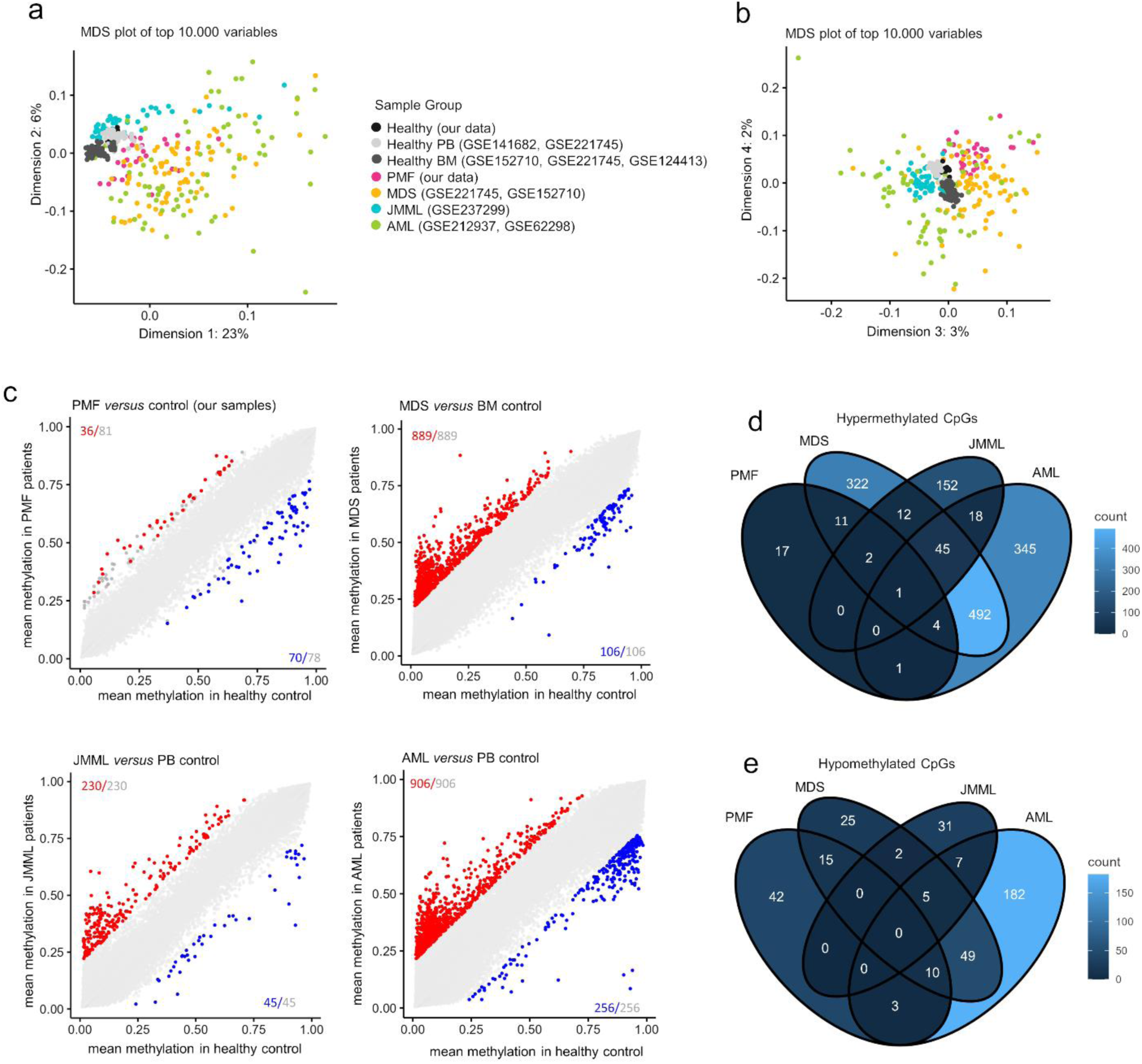
Comparison of myeloid malignancies after exclusion of cell-type specific CpGs. **a,b)** Multidimensional scaling plots of DNAm profiles (216,532 CpG sites) in patients with PMF, MDS, JMML, AML, and healthy controls (peripheral blood (PB) and bone marrow (BM)): a) first *versus* second dimension; a) third *versus* fourth dimension. **c)** Scatter plots showing mean DNAm beta values of healthy control *versus* PMF, MDS, JMML, or AML, accounting for potential differences between peripheral blood and bone marrow by using appropriate control sets. Significant hypo- and hypermethylated CpGs are indicated in blue and red (mean DNAm difference >20%; adjusted p-values < 0.05). Grey numbers indicate all CpGs exceeding the mean DNAm difference >20%, irrespective of statistical significance. **d,e)** Venn diagrams illustrating CpGs that are overlapping d) hyper- or e) hypomethylated in the above mentioned comparisons of four myeloid malignancies after the exclusion of cell-type specific CpGs.

Subsequently, we analyzed pairwise comparisons of DNAm changes between individual diseases and compounding controls (either BM or PB). Without filtering for cell type specific CpGs, we identified many significant CpGs for each disease (adj p < 0.05; difference of mean DNAm > 20%; Supplemental Figure 6c), and there was a considerable overlap in CpGs that were hyper- or hypomethylated across PMF, MDS, JMML, and AML (Supplemental Figure 6d-e). However, when we excluded CpGs with high variability between cell types, the differential DNAm signatures for each disease became significantly reduced and overlap among myeloid malignancies was minimal (Figure 6c-e). Only one CpG site (cg04470072_TC11) was hypermethylated across all these malignancies. Thus, while the DNAm changes in comparison to healthy blood samples can be largely attributed to the cellular composition, certain epigenetic aberrations may be indicative of specific diseases.

### Establishing an epigenetic signature for PMF

To determine if PMF possesses unique DNAm patterns that could aid in diagnosis, we performed pairwise comparisons between PMF and other myeloid malignancies, identifying significant differences in 387 CpGs for MDS, 700 for JMML, and 689 for AML (Supplemental Figure 7a-c). Among these, 17 CpGs were overlapping hypermethylated and 36 hypomethylated which are specific for PMF (Supplemental Figure 7d-e). Next, we analyzed if these CpGs were also differentially methylated in PMF *versus* heathy controls (Figure 6d-e). In fact, six hypomethylated CpGs (none of the hypermethylated CpGs) were overlapping in these comparisons: cg02210934 (no gene), cg02739280 (*NAV2*), cg21708058 (*TACC1*), cg07197092 (no gene), cg08069247 (*HABP2*), and cg04902833 (*C17orf99;* Supplemental Figure 8a). However, none of these CpGs could reliably discern all PMF from other samples. Furthermore, cg04902833 (*C17orf99*) was excluded from further analysis, because it was also hypomethylated in our control samples, indicating that it might be affected by batch effects or microarray versions.

Since single CpG analysis was insufficient for effective differentiation, we combined the remaining five CpGs into a PMF score (calculated as 5 minus the sum of the beta values, with higher scores indicating a stronger association with PMF). This score effectively distinguished PMF samples from all controls and most myeloid malignancies (Figure 7a). To further validate the score, we utilized other available datasets of PMF (16, 33), secondary myelofibrosis (16), ET (34), AML (35), pediatric AML (36), additional healthy controls, and own yet unpublished 450k profiles of post PV-MF, PV, and healthy controls (Supplemental Table S3). All five CpGs of the PMF-score showed clear offsets in MPN samples, except for the ET samples (Supplemental Figure 8b). The PMF-score was higher (>1) in almost all PMF, MF and PV samples (Figure 7b). Importantly, the PMF score did not correlate with the allele burden of driver mutations, reinforcing our earlier finding that epigenetic aberrations are heterogeneous within the malignant clone (Figure 7c). Furthermore, PMF-score did not reveal significant differences in our samples with *JAK2* V617F, *CALR*, and *MPL* mutation. Notably, in comparison to a public dataset of triple negative (TN) PMF samples, the PMF-score was lower in TN than in *JAK2* V617F or *CALR* mutated samples (Figure 7d).

**Figure 7:**
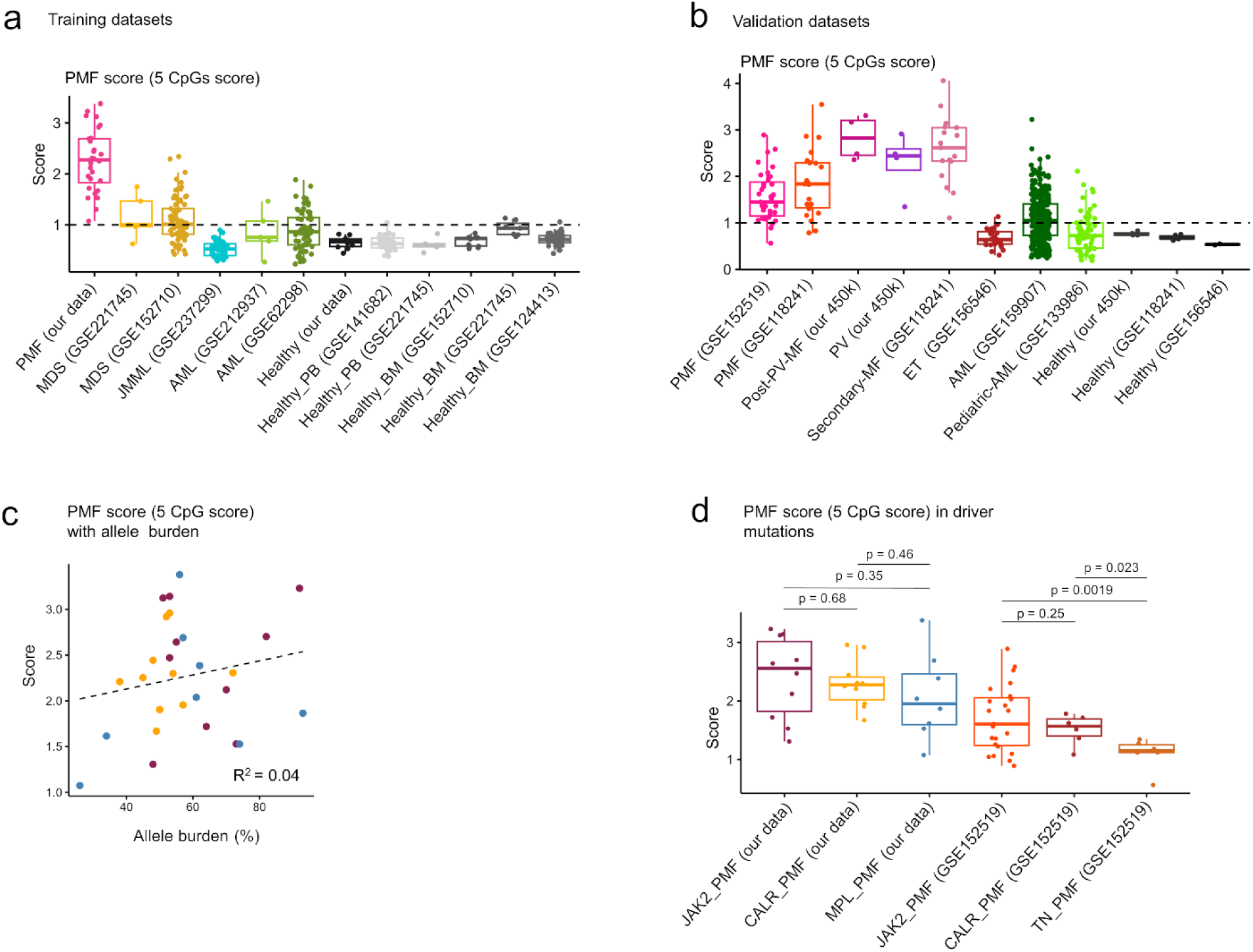
A five-CpG signature can discern PMF from other malignancies and controls. **a,b)** The DNAm levels at five CpGs (cg02210934, cg02739280, cg21708058, cg07197092, and cg08069247) were combined into a simple PMF-score (PMF-score = 5 - sum of the five DNAm values). The PMF-score is provided for datasets of the a) training set, and b) independent validation set. The datasets of PMF, other myeloid malignancies and healthy samples used from both our and public datasets are indicated with GSE numbers. **c)** The PMF score did not correlate with mutation allele burden. **d)** Box plot demonstrating that the driver mutations did not have significant impact on the PMF score. However, triple negative (TN) samples of publicly available PMF datasets (GSE152519) revealed a lower PMF-score. Statistical significance was determined using an unpaired t-test.

## Discussion

The results of this study indicate that the driver mutations in PMF have surprisingly little impact on the disease-associated epigenetic modifications. Significant DNAm differences between PMF with *JAK2* and *CALR* mutations were scarce, consistent with a recent study in ET patients (34). In contrast, MPL mutations appeared to lead to a slightly distinct epigenetic profile. The molecular link between the genomic and epigenetic modifications remains unclear. All three driver mutations activate the JAK-STAT signaling pathway, and while *JAK2* and *CALR* mutations were suggested to cause distinct mitotic defects leading to chromosomal instability (37), *MPL* mutation directly affect the structure and function of thrombopoietin receptor (38, 39).

When we investigated the impact of the *JAK2* V617F mutation in iPSCs, we observed no significant epigenetic changes. The moderate association of *JAK2* V617F PMF-associated DNAm changes in iHPCs might also be attributed to the pathognomonically skewed megakaryocytic and erythroid differentiation of iPSCs with the *JAK2* V617F mutation (20, 21). Thus, the iPSC model may not be ideally suited to recapitulate the complex epigenetic modifications that arise over many years during development of the disease. Additionally, the PMF-associated DNAm changes did not correlate with mutational allele burden, indicating that the epigenetic modifications are not homogenous in the entire malignant clone. Only very few CpGs correlated with allele burden, and it might be speculated that these arise earlier after the driver mutation and hence better reflect fraction of the malignant clone. Furthermore, the epigenetic makeup can also be altered in the non-malignant hematopoietic compartments or in the bone marrow microenvironment (40).

Previous studies suggested that alterations in the DNAm landscape play an important role in the pathogenesis and leukemic transformation of MPN (6). This might be caused by additional mutations in epigenetic writers, such as *ASXL1, DNMT3A*, *SRSF2*, and *TET2* (39, 41–43). Either way, the PMF-associated epigenetic changes were observed across different secondary mutations and many of these mutations are also frequently observed in other malignancies. This suggests that secondary mutations in epigenetic writers are not the sole drivers of the epigenetic changes seen in PMF.

Given that different cell types exhibit distinct DNAm profiles, it is crucial to consider cellular heterogeneity when interpreting DNAm data in hematological diseases (44). We excluded CpGs that displayed variability among different leukocyte subsets, demonstrating that previously noted differences in comparisons of diseased *versus* healthy blood largely stem from cellular composition. Furthermore, epigenetic aberrations have hardly been compared across different myeloid malignancies (45–47). Our integrative analysis across multiple studies demonstrated that also such comparisons are largely affected by differences in the cellular composition.

None of the individual CpGs could reliably discern PMF from healthy controls as well as from other myeloid malignancies, underscoring the need to combine multiple CpGs into comprehensive epigenetic signatures. Our 5 CpG PMF score offers a targeted analysis approach, compatible with methods such as pyrosequencing, BA-seq, or digital PCR, facilitating faster and more cost-effective assessments for clinical application (48). However, the PMF-score is not exclusive to PMF, as similar changes were also observed in PV, but not ET samples. Further investigation is needed to understand whether this is due to variations between studies, differences in allele burden, the duration of disease progression, or distinct pathophysiological mechanisms across various MPN entities. Additionally, it remains unclear whether AML and MDS samples with elevated PMF scores exhibit features of myelofibrosis.

In summary, the results of this study demonstrate that epigenetic patterns can differentiate myeloid malignancies, with the observed differences not directly linked to specific driver mutations but rather influenced by cellular composition and overlapping across various myeloid diseases.

## Further Information

### Data availability

All raw data are available from the corresponding authors upon reasonable request.

### Author Contributions

E.D.T. analyzed DNAm and gene expression data. V.T. performed iPSC experiments and supported analysis of DNAm data. K.K. and J.S. performed clinical data curation, data extraction, and data analysis. M.C. and S.K. conducted treatment and consenting of patients, logistics of bio samples, clinical interpretation, and data analysis. M.Z. provided the *JAK2* V617F iPSC lines and support in differentiation. W.W. initiated the research, designed, and supervised the study. V.T., E.D.T., and W.W. wrote the manuscript, and all authors approved the final version.

### Conflicts of Interest

W.W. and V.T. are involved in the company Cygenia GmbH (www.cygenia.com) that can provide service for epigenetic analysis to other scientists. Apart from this the authors have no competing interests to declare.

### Funding

This research was supported by funds from the German Research Foundation (Deutsche Forschungsgemeinschaft, DFG) within CRU344/417911533 “Untangling and Targeting Mechanisms of Myelofibrosis in Myeloproliferative Neoplasms” (W.W., S.K.), 363055819/GRK2415 (W.W.); WA 1706/12-2 (W.W.); WA1706/14-1 (W.W.); ZE 432/10-1 (M.Z.); KO2155/7-1 (S.K.), KO2155/7-2 (S.K.), KO2155/9-2 (S.K) and KO2155/6-1 (S.K); and the ForTra gGmbH für Forschungstransfer der Else Kröner-Fresenius-Stiftung (W.W.).

## Notes

### Author Declarations

Ethics committee of RWTH Aachen University Medical School gave ethical approval to this work.

