## Supplemental Methods Figures and Tables for "DNA methylation in primary myelofibrosis is partly associated with driver mutations and distinct from other myeloid malignancies"

#### Preprocessing and further analysis of DNA methylation data

Analysis of the DNAm data was performed in R (4.4.0) (1). Initial quality control of the DNA methylation data was performed using the minfi package (version 1.50.0). Raw data (.IDAT files) of both datasets were independently preprocessed using the SeSAMe package for R v1.22.2 (2), in the following order: quality masking the probes with poor design, reset the color channel for Type-I probes inference, non-linear dye bias correction, detection of p-value using out-of-band (OOB) array hybridization, and background correction. Additionally, CpGs at the X and Y chromosomes were removed. Probes that failed in 10% or more of our samples were excluded and resulting 812,274 CpGs. The limma R package (3.60.0) was used for calculation of Benjamini-Hochberg adjusted p values and the multidimensional scaling plots. Relevant DNAm changes were defined as showing at least 20 % difference in mean beta values and an adjusted p value ≤ 0.05. For gene ontology (GO) analysis the R package missMethyl (1.38.0) was used. The R packages: watermelon, ggplot2, ggrepel, ggbeeswarm, reshape2, ggExtra, gprofiler2, ComplexHeatmap, and VennDiagram were used for graphical presentation.

DNAm analysis of the raw data from iPSCs and their hematopoietic derived cell samples was analyzed as described as above, and the probes with failed detection P values in three or more samples were excluded, resulting in 630,700 CpGs. Furthermore, we only considered CpGs that were represented by the EPICv1 and EPICv2 BeadChip platforms for the comparison analysis of patient derived samples and iPSC derived samples, with resulting in 590,349 CpGs.

#### Correlation with gene expression data

Gene expression analysis was performed on 9 PMF patient and 21 healthy control samples from publicly available GSE26049 dataset. Raw CEL files were processed in R using the affy R package (1.82.0), and initial quality assessment was performed to evaluate data quality. The Robust Multi-array Average (RMA) algorithm was applied for background correction, normalization and probe set level expression values calculation. Differential expression analysis was performed with the limma package (3.60.0), applying linear models to compare expression levels between PMF patients and healthy controls, and adjusting for multiple testing to control the false discovery rate (FDR). Probes with a fold change greater than 1 and adjusted p value is less than 0.05 were identified as significantly differentially expressed. Differentially expressed genes were annotated using Affymetrix Human Genome U133 Plus 2.0 Array annotation file and genes that are significantly expressed are selected for future analysis.

#### Generation of JAK2 V617F iPSC and their hematopoietic lineage differentiation

Induced pluripotent stem cells (iPSC) were generated from PBMCs of three PV patients with varying *JAK2* V617F allele burdens (37%, 96%, and 25%, respectively) in a previous work, as described before (3-5). Using CRISPR/Cas9 genome editing, additional genotypes were generated for each patient when required to generate the full complement of wild type, heterozygous, and homozygous clones. In the Human Pluripotent Stem Cell Registry, these iPSCs are listed as: patient 1 WT (UKAi002-A), heterozygous (UKAi002-B) and homozygous *JAK2* V617F clone (UKAi002-B3); patient 2 WT (UKAi003-A1), heterozygous (UKAi003-A2) and homozygous clones (UKAi003-A); patient 3 WT (UKAi013-A), heterozygous (UKAi013-B) and homozygous clones (UKAi013-B1). iPSCs were routinely maintained in StemMACS iPS-Brew XF on 6-well plates coated with vitronectin.

Hematopoietic differentiation of iPSC clones was performed as described previously (6). In brief, iPSCs were cultured in micro-contact printed plates with StemMACS iPS Brew XF medium (Miltenyi Biotec) and 10 µM Y-27632 (Abcam, Cambridge, United Kingdom) to form embryonic bodies (EB) (7). Self-detachment of EBs was observed after 6 to 9 days, depending on the clone, and they were then resuspended in serum-free medium containing 50% IMDM, 50% Ham's F12, 1% chemically defined lipid concentrate, 2 mM GlutaMAX (all Thermo Fisher Scientific), 0.5% Albiomin (Unifols), 400 µM 1-thioglycerol, 50 µg/mL L-ascorbic acid, and 6 µg/mL holo transferrin (all Sigma Aldrich, St. Louis, MO, USA) supplemented with 10 ng/mL FGF-2 (Peprotech, Hamburg, Germany) and 10 ng/mL BMP-4 (Miltenyi Biotec). Approximately 30 to 50 EBs were distributed per well on a gelatin coated 6-well plate. From day 2 to day 7, cells were cultured in serum-free medium supplemented with 10 ng/mL FGF 2, 10 ng/mL BMP-4, 50 ng/mL SCF, 10 ng/mL VEGF-A (all Peprotech), and 10 U/mL penicillin/streptomycin

(Thermo Fisher Scientific). From day 8 to day 16, serum-free medium was supplemented with 10 ng/mL FGF 2 and 50 ng/mL SCF only. Cells were harvested on day 16 and their phenotype was analyzed by flow cytometry. Further cells were collected and isolated using QIAamp DNA Mini Kit (Qiagen) for DNA methylation analysis.

**DNA methylation analysis by targeted bisulfite amplicon sequencing (BA-seq)**

DNAm levels at the three age-associated CG dinucleotides (CpG sites) that are associated with the coiled-coil domain-containing protein 102B (*CCDC102B*), four and a half LIM domains protein 2 (*FHL2*), and phosphodiesterase 4C (*PDE4C*) were analyzed using targeted bisulfite amplicon sequencing (BA-seq), as described in detail before (8, 9). Genomic DNA from cryopreserved whole blood cells after red blood cells depletion and single CFU colonies was bisulfite converted with the EZ DNA Methylation Kit (Zymo Research, Irvine, USA) and amplified using primers with handle sequences. All three amplicons for each donor were pooled for barcoded second PCR, then subsequently amplified with Illumina adapters and eventually subjected to 250 bp pair-end sequencing on a MiSeq lane using the Miseq reagent V2 Nanokit (both from Illumina). FastQ files from MiSeq analysis were aligned to the reference genome hg19 using the Bismark tool (10) and DNA methylation values determined with the Bismark methylation extractor. For heatmaps, the frequencies of DNA methylation patterns in individual reads were calculated by the number of reads containing the pattern divided by the total number of reads of the target region per sample. The most abundant reads of similar patterns within neighbouring CpGs were grouped for visualization with Python's package seaborn (11).

**Colony forming unit assay**

Peripheral blood mononuclear cells from MPN and healthy donors were cultured in a semisolid medium for 14 days to perform CFU assay with $1x10^6$ cells per condition. Colonies were counted, classified, and DNA was isolated using the NucleoSpin XS Tissue Kit (Macherey-Nagel, Düren, Germany) to determine the mutation status of *JAK2* V617F mutation, as previously described (12).

### Supplemental Figures

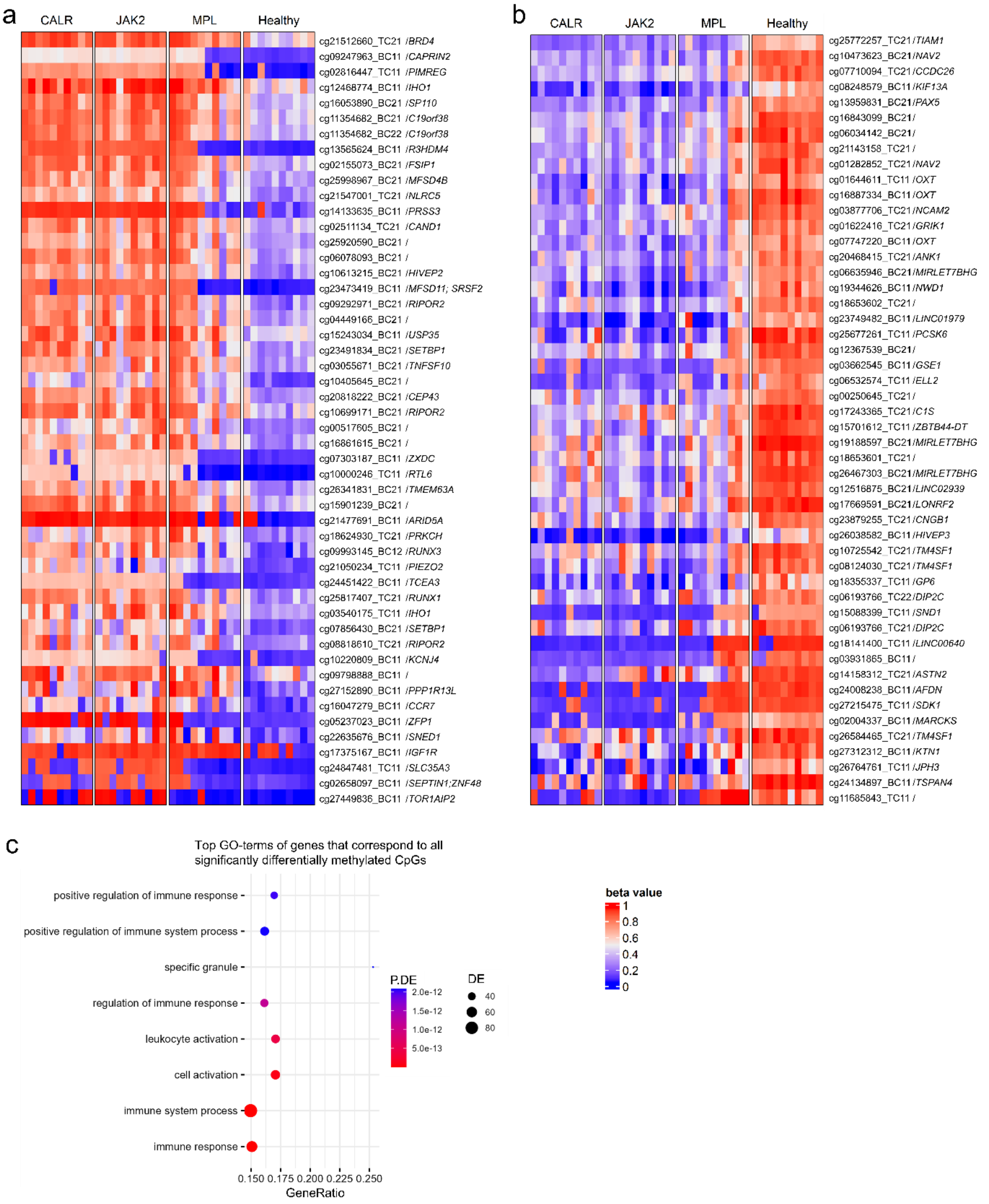

**Figure S1: DNA methylation profile of PMF patients differs from healthy controls.**

**a,b)** Heatmap of top 50 a) hypermethylated and b) hypomethylated CpG sites in PMF *versus* healthy regardless of driver mutation difference. **c)** Gene Ontology classification was performed on significantly differentially methylated CpGs in PMF patients compared to healthy controls. Terms with an FDR below 0.05 are provided. The gene ratio is the proportion of differentially expressed genes relative to the total number of genes in the set, with DE representing differentially methylated genes and P.DE indicating the p-value for the GO term's overrepresentation.

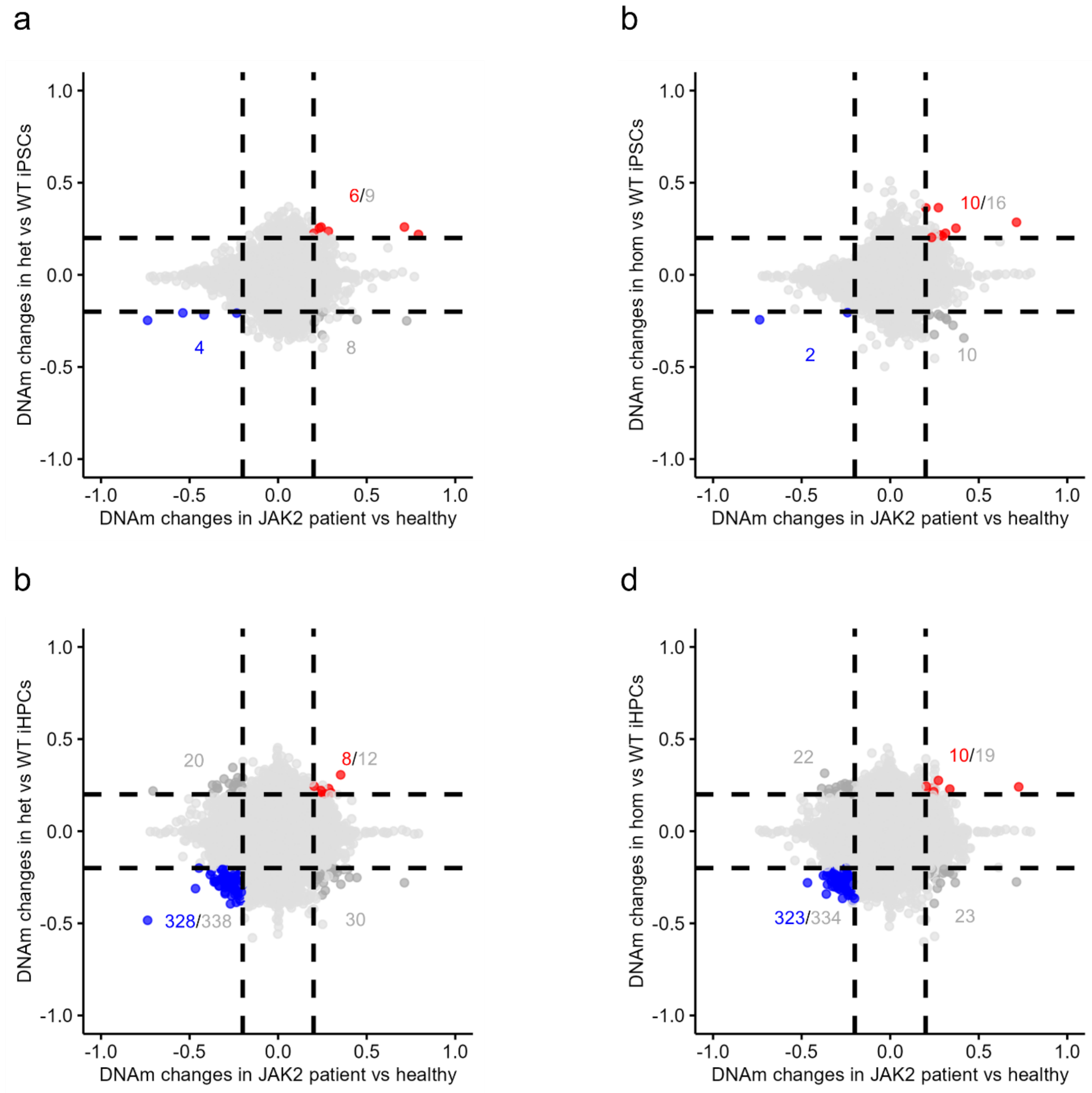

**Figure S2: Association of *JAK2* V617F-associated DNAm in patients with the iPSC model.**
Comparison of significantly differentially methylated CpGs between *JAK2* patient and healthy control with **a)** heterozygous iPSCs *vs*. WT iPSCs, **b)** homozygous iPSCs *vs*. WT iPSCs, **c)** heterozygous iPSC-derived hematopoietic progenitor cells (iHPCs) *vs*. WT iHPCs, and **d)** homozygous iHPCs *vs*. WT iHPCs. CpGs that are hypomethylated or hypermethylated in both groups are indicated in blue and red, respectively.

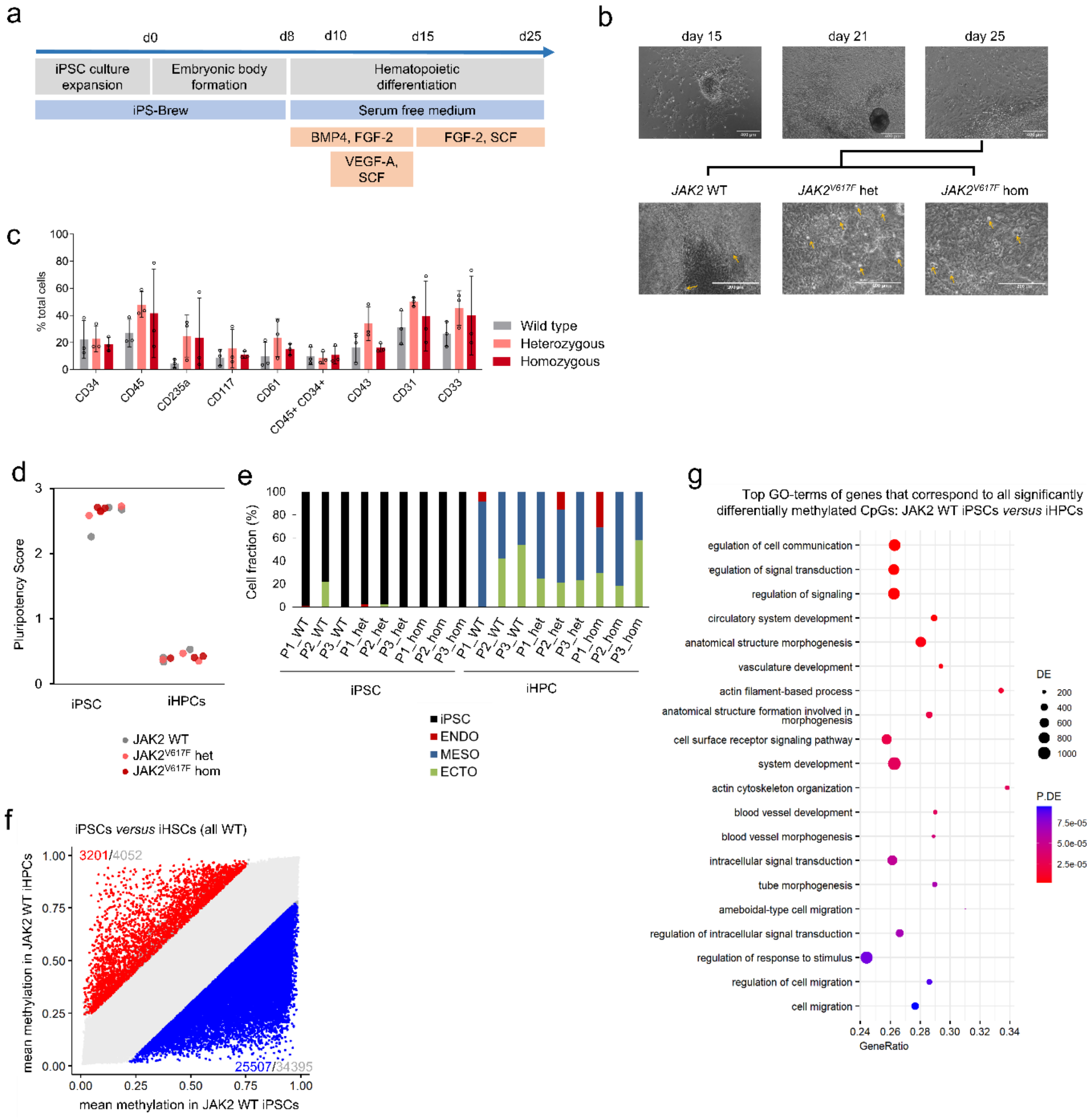

**Figure S3: iPSCs differentiation into hematopoietic stem and progenitor cells.**

**a)** Hematopoietic differentiation protocol. **b)** Exemplary phase contrast image of iPSC derived hematopoietic differentiated cells (iHPCs) during different days of differentiation for all three genotypes: WT, heterozygous (het) and homozygous (hom) *JAK2* V617F mutant. Hematopoietic stem cells emerged from the hemogenic endothelium niche which formed a typical cobblestone structure (yellow arrows). **c)** Flow cytometric analysis of hematopoietic surface markers on harvested iHPCs at day 16 of differentiation (n = 3 for each genotype). **d)** iPSCs and iHPCs were compared for PluripotencyScore and **e)** to track the differentiation of the three germ layers (13). **f)** DNAm profiles of WT iPSCs and iHPCs were compared to identify differentially methylated CpGs. The numbers indicate the difference in mean methylation of the cut-off 0.2 in grey and those that are significant in red (hypermethylated) or blue (hypomethylated). **g)** Gene Ontology analysis of the significant CpGs with a false discovery rate (FDR) of less than 0.05 that were differentially methylated in both hypo- and hypermethylated genes when comparing WT iHPCs with iPSCs.

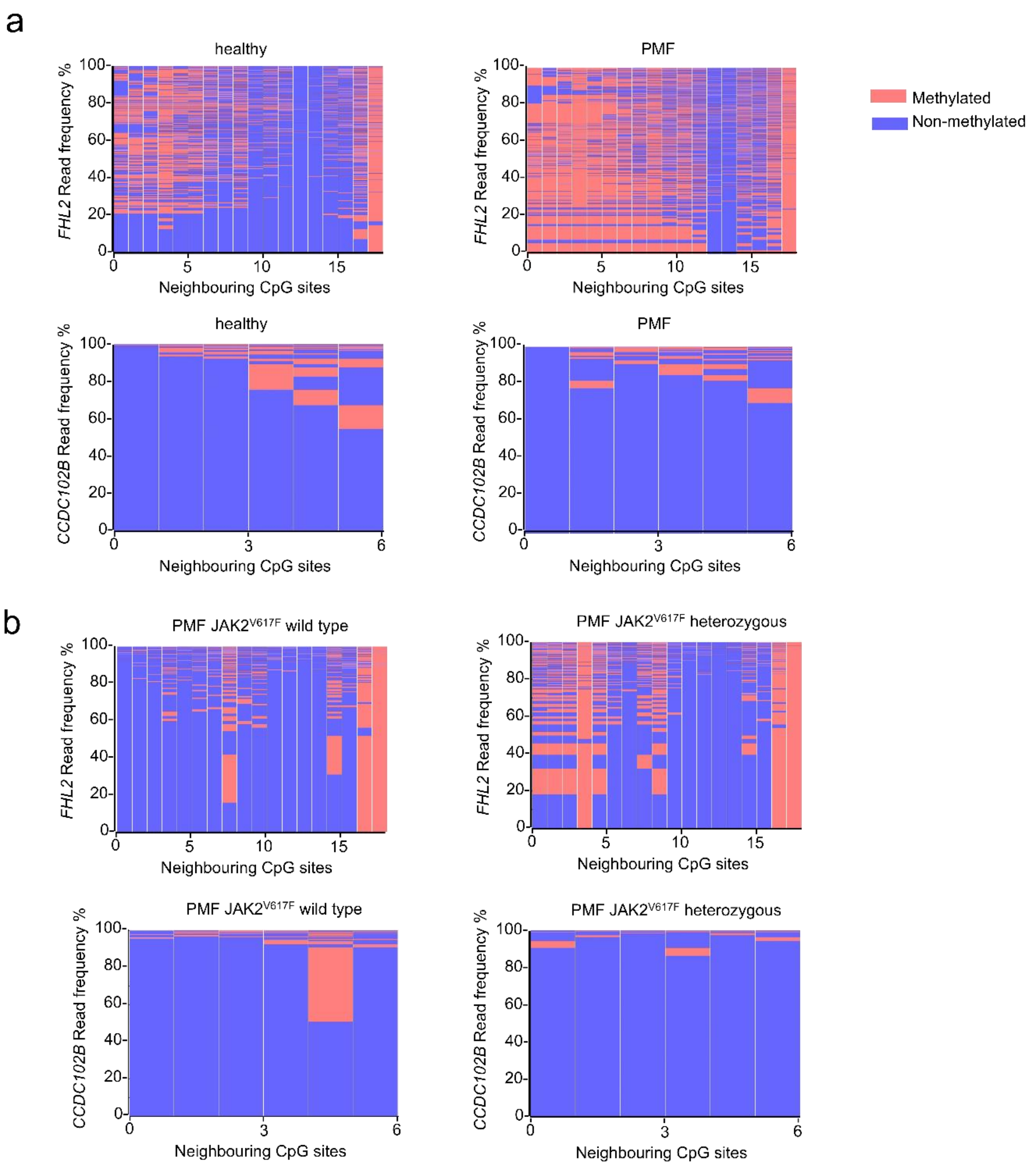

**Figure S4: Heterogeneity of epigenetic age predictions based on individual amplicon reads.**

**a)** Heatmap depicts exemplarily the frequencies of DNAm patterns within the neighbouring CpGs of the *FHL2* amplicon and for the *CCDC102B* amplicon in BA-seq data of a healthy donor, and a PMF patient of the same age (in analogy to figure 4 e,f). In healthy and MPN samples the patterns of neighbouring CpGs are quite heterogeneous and there is no obvious clonal DNAm pattern in the patient samples. **b)** Individual colony forming units (CFUs) were analyzed after 14 days (in analogy to figure 4 g,h). The heat maps exemplarily depict the frequencies of DNAm patterns within the neighbouring CpGs of the *FHL2* and *CCDC102B* amplicon in BA-seq data of wild type, and *JAK2* V617F mutated colonies from the same patient. In comparison to the corresponding heatmaps of blood there are prominent patterns in individual colonies, which might reflect the pattern of the initial colony forming cell.

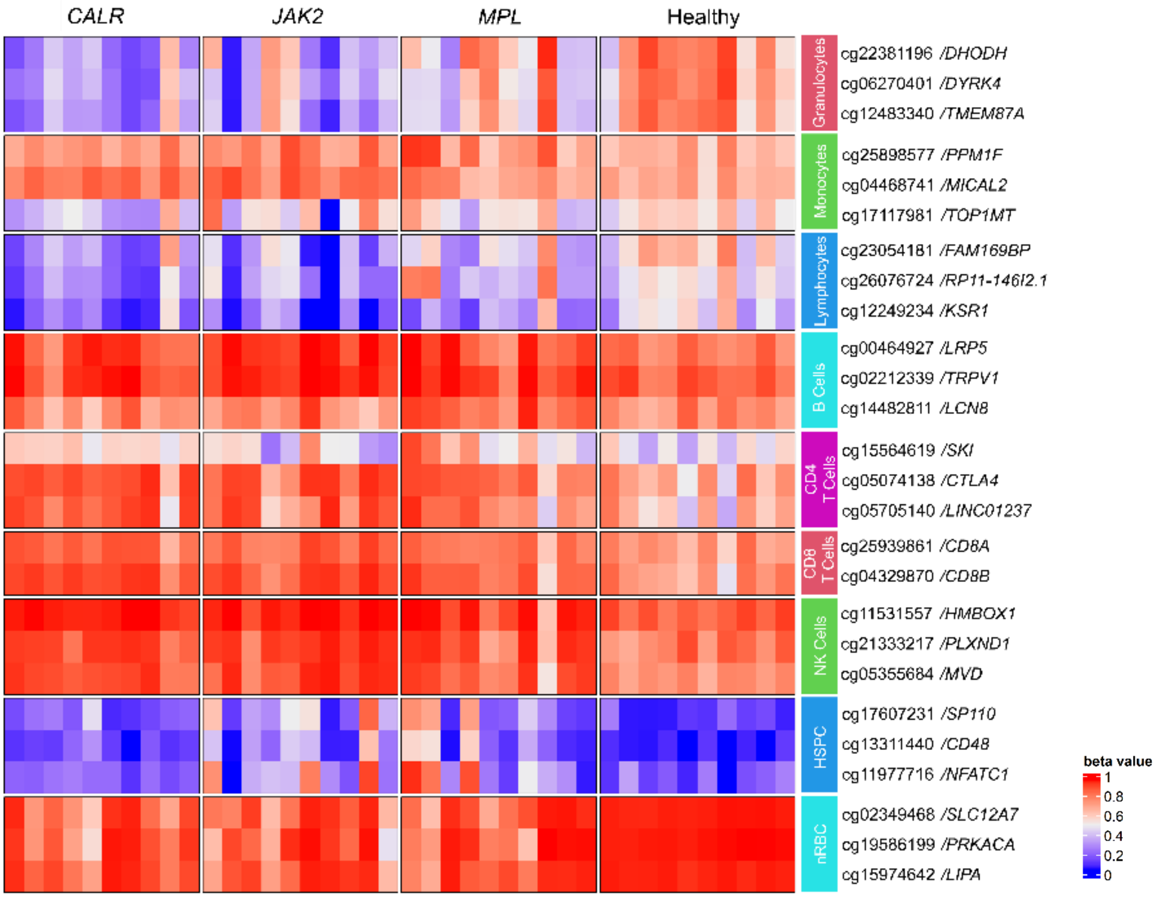

**Figure S5: Heatmap of DNAm at 26 cell-type specific CpG sites.**

The heatmap illustrates DNAm (beta values) in PMF and control samples focussing on the cell type specific CpG sites identified before (14). Granulocytes exhibited more pronounced hypomethylation in PMF samples, consistent with the anticipated higher proportion of granulocytes in these samples.

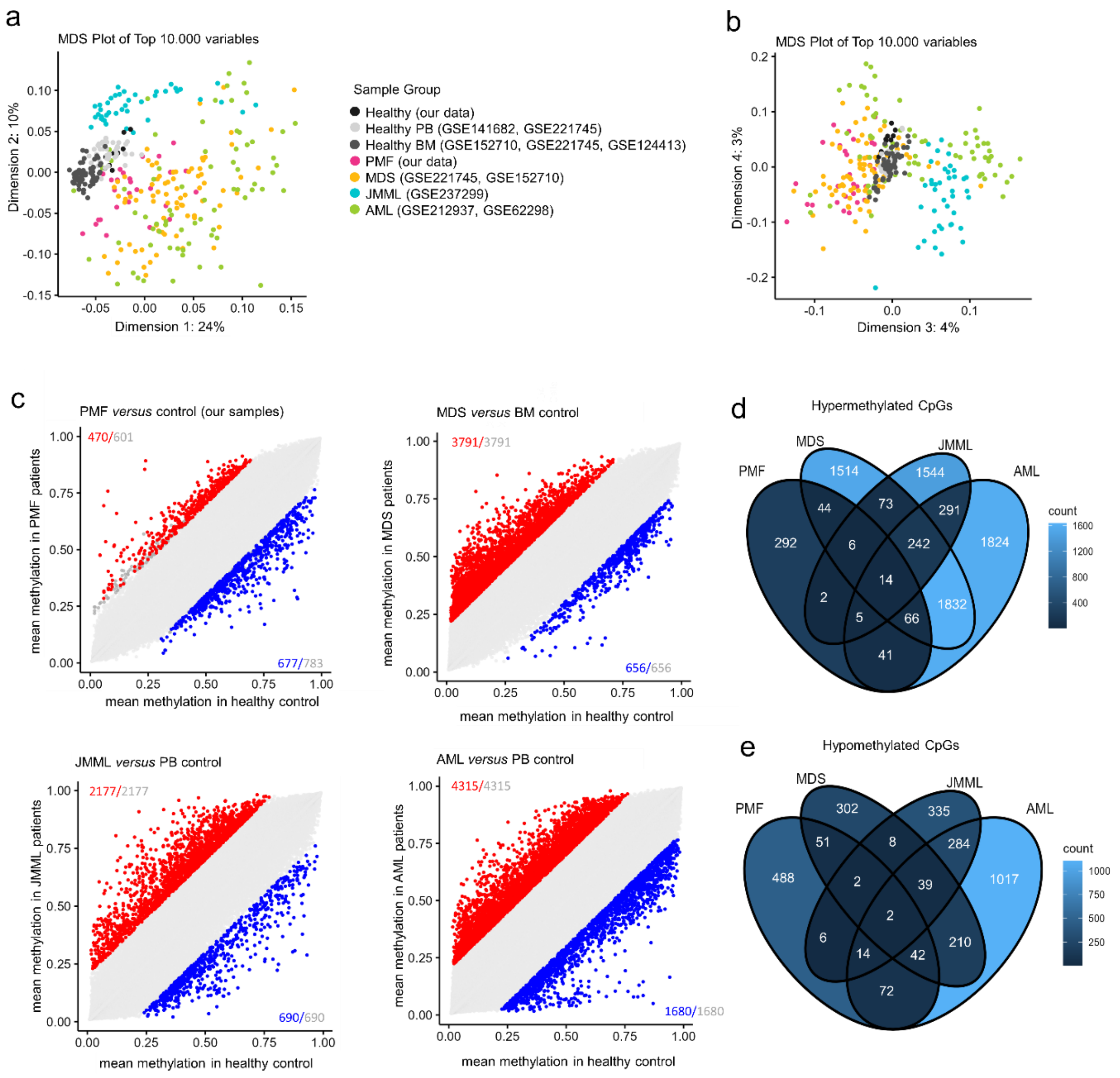

**Figure S6: Comparison myeloid malignancies with healthy controls.**

**a,b)** The multidimensional scaling plots presents DNA methylation profiles (304,145 CpG sites) from patients with PMF, three other myeloid malignancies (MDS, JMML, AML), and healthy controls, based on whole blood and bone marrow samples. The plot shows a) the 1st and 2nd dimension and b) the 3rd and 4th dimension. **c)** Scatter plots compare mean DNAm beta values from healthy controls with PMF, MDS, JMML, and AML samples. The same tissue type (PB or BM) of healthy sample was used for comparison. **d,e)** Venn diagrams illustrate CpGs in all myeloid malignancies compared to healthy controls for d) hypermethylated and e) hypomethylated CpGs. A total of 14 hypermethylated CpGs and 2 hypomethylated CpGs were found to overlap across all myeloid malignancies. Information on these CpGs is provided in the supplemental table S6.

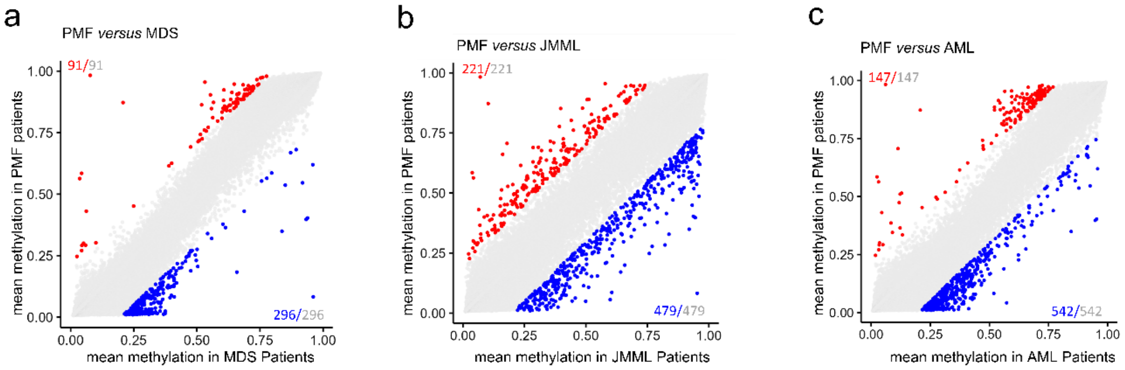

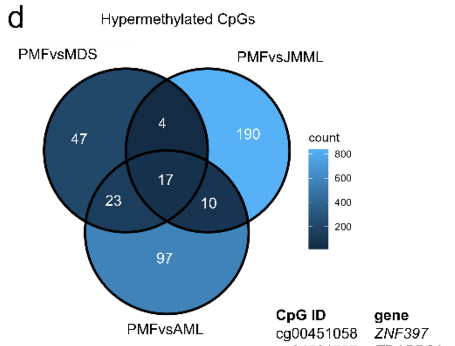

| CpG ID | gene |
|---|---|
| cg00451058 | *ZNF397* |
| cg24521757 | *TRAPPC9* |
| cg01957330 | *SKAP2* |
| cg02159896 | *MPPE1* |
| cg25568561 | *PPP2R5C* |
| cg22986271 | *SACM1L* |
| cg17004373 | *GNL2* |
| cg18251019 | *OBFC1* |
| cg04891577 | *CYFIP1* |
| cg10985479 | *LEF1* |
| cg09036531 | |
| cg13810766 | *PRKAG2* |
| cg09190325 | *HMGN4* |
| cg04205664 | *CLEC2L* |
| cg14497545 | *MAML3* |
| cg03027241 | *KCNG1* |
| cg24724689 | |

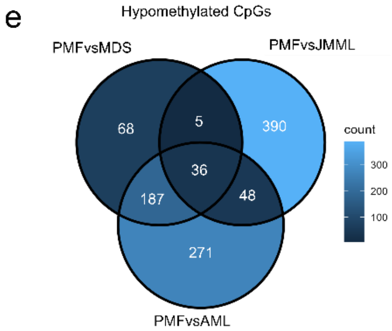

| CpG ID | gene |
|---|---|
| cg04560503 | *TRRAP* |
| cg04902833 | *C17orf99* |
| cg05788548 | *SKI* |
| cg26127925 | |
| cg02488871 | *VIL1* |
| cg27292870 | |
| cg04626451 | *PHACTR2* |
| cg04683551 | *HSPA14* |
| cg23215407 | |
| cg16907029 | *KCNA1* |
| cg21708058 | *TACC1* |
| cg12026095 | *FTL* |
| cg01684248 | |
| cg04450857 | *EMX2OS* |
| cg13003239 | *WDR6* |
| cg18210026 | |
| cg18141888 | |
| cg10853231 | *GLTSCR1* |
| cg02899001 | |
| cg25152348 | *NCAPH2* |
| cg08206623 | *CDKN1C* |
| cg01204305 | *AMIGO3* |
| cg07197092 | |
| cg12457901 | *VARS2* |
| cg08069247 | *HABP2* |
| cg02739280 | *NAV2* |
| cg02210934 | |
| cg27639620 | *TSPAN4* |
| cg21383487 | |
| cg01745789 | |
| cg11637712 | *ARHGAP23* |
| cg23903465 | *FAM65B* |
| cg06636971 | *FAM65B* |
| cg08576643 | *FAM65B* |
| cg26922444 | *FAM65B* |
| cg26764761 | *JPH3* |

**Figure S7: Pairwise comparison of PMF *versus* other myeloid malignancies.**

**a-c)** Scatter plots of the mean DNAm beta values of PMF patients *versus* a) MDS patients, b) JMML patients, c) AML patients. **d,e)** Venn diagram depict overlapping CpGs identified by pairwise comparisons of PMF with other malignancies for d) hypermethylated and e) hypomethylated CpGs. For the overlapping CpGs the IDs and corresponding gene names are provided.

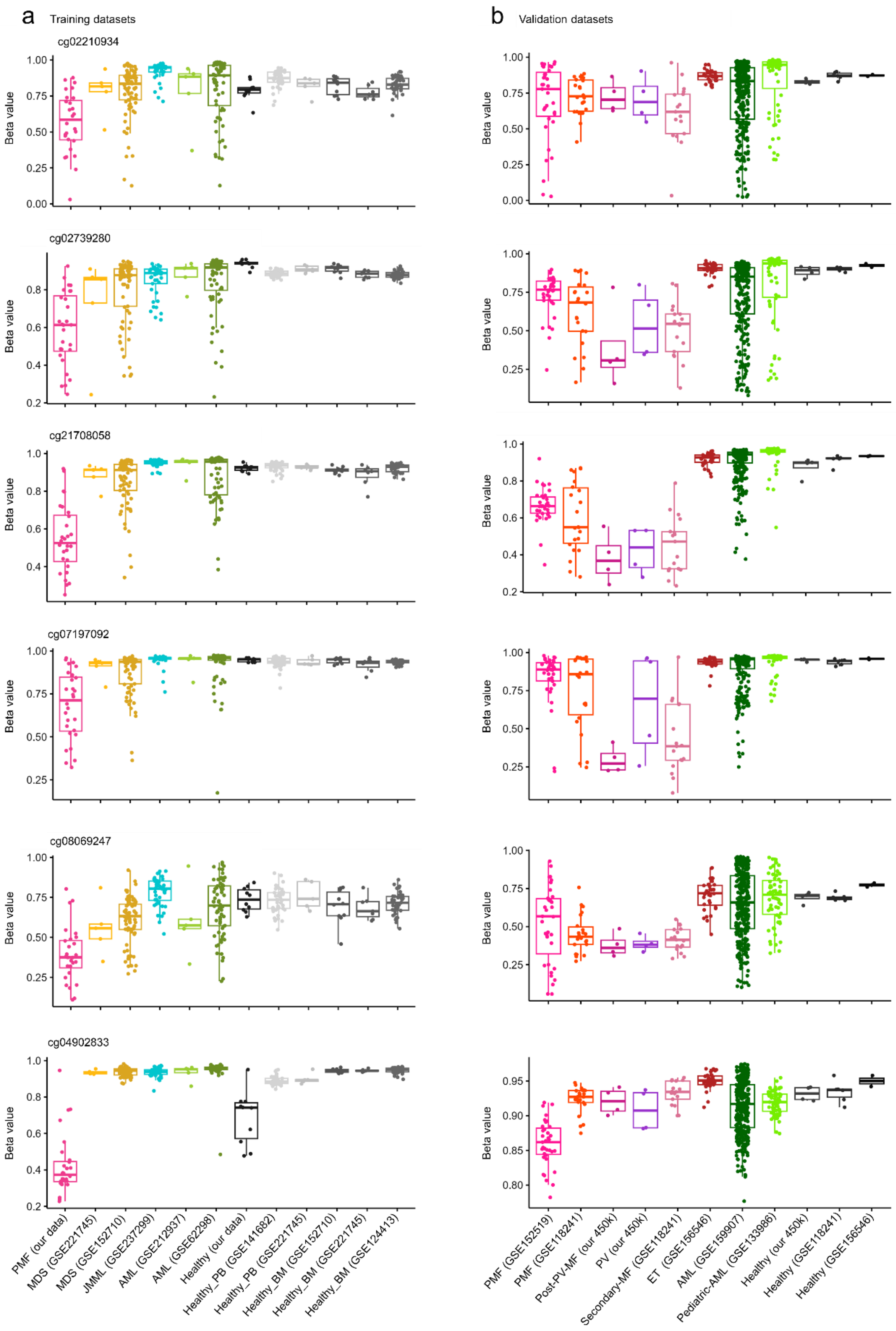

**Figure S8: Analysis of six PMF-associated CpGs across different datasets.**

PMF-specific CpGs identified from pairwise comparisons of all myeloid malignancies with healthy samples (Figure 6 d,e) were compared with the overlapping CpGs derived from pairwise comparisons of all malignancies (Figure S7d,e). 6 hypomethylated CpGs were found to be overlapping. Box plot displays the DNAm values of these CpGs **a)** in training datasets and **b)** in validation dataset. Since cg04902833 exhibited hypomethylation in healthy samples as well, it was not considered for the PMF score.

### Supplemental Tables

**Supplemental Table S1. Information on MPN samples.**

| Sample Number | Gender | Disease | Driver Mutation | Allele Burden | Age range | Additional mutations | Therapy |
|---|---|---|---|---|---|---|---|
| **1** | Female | PMF | CALR | 72% | 56-60 | | w&w |
| **2** | Male | PMF | CALR | 57% | 51-55 | | w&w |
| **3** | Female | PMF | CALR | 54% | 56-60 | *ASXL1* | Ruxolitinib |
| **4** | Male | PMF | CALR | 53% | 61-65 | | Momelotinib |
| **5** | Female | PMF | CALR | 52% | 61-65 | | Ruxolitinib |
| **6** | Male | PMF | CALR | 50% | 26-30 | | w&w |
| **7** | Female | PMF | CALR | 49% | 21-25 | | w&w |
| **8** | Male | PMF | CALR | 48% | 71-75 | | w&w |
| **9** | Female | PMF | CALR | 45% | 51-55 | *ASXL1* | w&w |
| **10** | Female | PMF | CALR | 38% | 56-60 | | Ruxolitinib |
| **11** | Female | PMF | JAK2 | 92% | 66-70 | *TET2 1 bp Del, TET2 2 bp Del* | Momelotinib |
| **12** | Male | PMF | JAK2 | 82% | 56-60 | | Ruxolitinib |
| **13** | Male | PMF | JAK2 | 73% | 76-80 | *ASXL1, SRSF2 P95L* | w&w |
| **14** | Female | PMF | JAK2 | 70% | 51-55 | *SRSF2 24bp Del* | w&w |
| **15** | Male | PMF | JAK2 | 64% | 66-70 | | w&w |
| **16** | Female | PMF | JAK2 | 55% | 76-80 | *ASXL1* | Ruxolitinib, Hydroxyurea |
| **17** | Female | PMF | JAK2 | 53% | 71-75 | *DNMT3A* | Hydroxyurea |
| **18** | Female | PMF | JAK2 | 53% | 66-70 | | w&w |
| **19** | Male | PMF | JAK2 | 51% | 61-65 | *ASXL1* | Ruxolitinib |
| **20** | Male | PMF | JAK2 | 48% | 61-65 | | Ruxolitinib |
| **21** | Male | PMF | MPL | 93% | 71-75 | *CALR 52bp Del, SRSF2 P95L* | Ruxolitinib |
| **22** | Male | PMF | MPL | 74% | 71-75 | *IDH2 R140Q, SRSF2 P95R, SRSF2 R186L* | Ruxolitinib |
| **23** | Male | PMF | MPL | 62% | 76-80 | | w&w |
| **24** | Male | PMF | MPL | 57% | 86-90 | *SRSF2 3bp Dup, TET2 6bp Del, TET2 1bp Del* | w&w |
| **25** | Male | PMF | MPL | 78% | 86-90 | *NFE2, TET2 R552* | w&w |
| **26** | Male | PMF | MPL | 34% | 56-60 | | w&w |
| **27** | Male | PMF | MPL | 56% | 61-65 | *ASXL1, TET2* | w&w |
| **28** | Male | Pre-PMF | MPL | 26% | 76-80 | | Hydroxyurea |
| **29** | Male | ET | MPL | 38% | 51-55 | | Ruxolitinib |
| **30** | Female | ET | MPL | 39% | 66-70 | | w&w |

w&w = watch and wait

**Supplemental Table S2: Datasets of sorted leukocyte subsets.**

This table is provided as a separate Excel file. It includes sample descriptions (GSE, GSM, GEO title), array-related information, cell type group, and if available, details on age, gender, and isolation method.

**Supplemental Table S3: Information on the GEO datasets of training and validation sets.**

This table is provided as a separate Excel file. It includes sample descriptions (GSE, GSM, GEO title), array-related information, cell type group, if available, details on age, gender.

**Supplemental Table S4: Differentially methylated CpG between driver mutations.**

This table is provided as a separate Excel file. It contains separate sheets for 3 different pairwise comparisons: *JAK2* mutation *versus CALR* mutation, *JAK2* mutation *versus MPL* mutation, *CALR* mutation *versus MPL* mutation. Significantly hypermethylated CpGs are highlighted in red and significantly hypomethylated CpGs in blue.

**Supplemental Table S5: CpGs associated with allele burden.**

This table is provided as a separate Excel file. To determine how individual DNAm values correlate with allele burden, we calculated linear regressions and filtered CpGs with a slope >0.0035. These CpGs are sorted by Pearson correlation. CpG names are provided along with associated annotation information (chr, gene symbol, position, regulatory feature group).

**Supplemental Table S6: Overlapping CpGs in myeloid malignancies *versus* healthy controls.**

| Name | Diff mean_ PMF | Diff mean MDS | Diff mean_ JMML | Diff mean_ AML | Relation to_Island | UCSC RefGene Name | UCSC RefGene Group |
|---|---|---|---|---|---|---|---|
| cg00524708 | 0,22 | 0,32 | 0,30 | 0,31 | N_Shore | *PRSS36* | Body |
| **cg02676175** | **0,25** | **0,26** | **0,26** | **0,26** | **Island** | ***KCTD11; ACAP1*** | **TSS1500; Body** |
| cg02938205 | 0,29 | 0,56 | 0,28 | 0,56 | Island | *CCDC36* | TSS200;5'UTR |
| cg04470072 | 0,28 | 0,52 | 0,22 | 0,50 | Island | *PSKH2* | TSS200 |
| cg05491854 | 0,28 | 0,24 | 0,34 | 0,32 | N_Shore | *FAM65B* | 5'UTR |
| cg08818610 | 0,38 | 0,35 | 0,40 | 0,40 | Island | *FAM65B* | 5'UTR |
| cg11203616 | 0,20 | 0,33 | 0,25 | 0,27 | Island | *TNS3* | 5'UTR |
| cg12439325 | 0,20 | 0,29 | 0,21 | 0,22 | Island | *TMPRSS12* | 1stExon |
| cg12440062 | 0,28 | 0,31 | 0,21 | 0,22 | OpenSea | *CRISP2* | TSS200 |
| cg14095959 | 0,23 | 0,25 | 0,26 | 0,23 | Island | | |
| cg14301190 | 0,24 | 0,26 | 0,28 | 0,30 | Island | *PRSS36* | Body |
| cg15289427 | 0,25 | 0,38 | 0,37 | 0,52 | Island | *FAM65B* | 5'UTR |
| cg18675097 | 0,30 | 0,52 | 0,39 | 0,42 | Island | *NKAPL* | 5'UTR;1stExon |
| cg26398848 | 0,22 | 0,27 | 0,22 | 0,28 | Island | *ACP1* | TSS1500 |
| cg22879098 | -0,28 | -0,29 | -0,34 | -0,22 | OpenSea | TPPP | Body |
| cg24082121 | -0,25 | -0,36 | -0,37 | -0,23 | OpenSea | TPPP | Body |
